# Impact and Equality of Expanding Pre-Exposure Prophylaxis (PrEP) for Men who have Sex with Men in Los Angeles County

**DOI:** 10.1101/2021.09.03.21263101

**Authors:** Anthony Nguyen, Emmanuel Fulgence Drabo, Wendy Garland, Corrina Moucheraud, Ian W Holloway, Arleen Leibowitz, Sze-chuan Suen

## Abstract

**Background:** Racial and ethnic minority men who have sex with men (MSM) are disproportionately affected by HIV/AIDS in Los Angeles County (LAC), an important epicenter in the battle to end HIV.

**Objective:** To examine tradeoffs between effectiveness and equality of PrEP allocation strategies among different racial and ethnic groups of MSM in LAC.

**Design, Setting, and Population:** We developed a microsimulation model of HIV among MSM in LAC using county epidemic surveillance and survey data to capture demographic trends and subgroup-specific partnership patterns, disease progression, patterns of PrEP use, and patterns for viral suppression.

**Intervention:** We simulated interventions where an additional 3000, 6000, or 9000 PrEP prescriptions are provided annually in addition to current levels, following different allocation scenarios to each racial/ethnic group (Black, Hispanic, or White).

**Measurements:** We estimated cumulative infections averted and measures of equality, after 15 years (2021-2035), relative to base case (no intervention).

**Results:** Of the policies evaluated, targeting PrEP preferentially to Black individuals would result in the largest reductions in incidence and disparities. This outcome was robust to different partnership preference assumptions, though the magnitude of impact differs.

**Limitations:** We limit analysis to MSM, who bear the majority of HIV/AIDS burden in LAC. We do not consider transmission via injection drug use or mother-to-child transmission, nor do we capture individual network transmission effects. We assume no improvements in the prevention-diagnosis-treatment cascade besides increased PrEP use.

**Conclusions:** We find there is little trade-off between effectiveness and equality of outcome when choosing groups to target for PrEP in LAC – by focusing on MSM with the highest HIV incidence (Black), we can reduce both overall infections and racial/ethnic disparities.

## INTRODUCTION

### HIV in Los Angeles County

The HIV epidemic in Los Angeles County (LAC) remains one of the largest nationwide, with approximately 52,000 people living with HIV (PLWH) and over 1,600 new HIV diagnoses annually.^1–4^ Men who have sex with men (MSM) comprise 83% of the PLWH in LAC (compared to 61% nationally^5^).

There exist profound racial and ethnic disparities in HIV burden and care in LAC among MSM. An estimated 17.5% are non-Hispanic Black, although Black MSM represent only 7.9% the MSM population.^6^ While 80% of diagnosed non-Hispanic White MSM were linked to care within a month, only 65% and 73% of non-Hispanic Black and Hispanic MSM were linked.^6^ Similar trends are seen in engagement in HIV care and viral suppression.^6^ While non-Hispanic White MSM and Hispanic MSM have similar retention in care proportions, (60% and 61% respectively), the retention among non-Hispanic Black MSM is lower, at 52%.^6^

Developing county-specific HIV studies that account for these differences in HIV risk is critical to better understand and design strategies across population subgroups.

### Combatting the HIV Epidemic

The Ending the HIV Epidemic (EHE) initiative intends to end the HIV epidemic in the United States within ten years.^7^ It emphasizes four pillars, including preventing new infections through the use of pre-exposure prophylaxis (PrEP),^7,8^ a biomedical prevention strategy which can reduce HIV infection risk by up to 99%^8,9^ and is highly cost-effective.^10–13^ LAC aims to accelerate efforts that increase PrEP use, particularly for populations with high HIV diagnosis rates and low PrEP coverage, such as Black and Latino MSM, to reduce racial and ethnic disparities in HIV incidence.^14^

We therefore evaluated a variety of PrEP allocation strategies for MSM in LAC to determine their effectiveness in reducing new HIV infections and in narrowing racial and ethnic disparities in HIV incidence. Most studies to date have examined population-level effects, which mask potential disparities in outcomes for specific population subgroups.^10–12,15–18^ Notable exceptions include agent-based models of HIV transmission among MSM in Baltimore, MD^16^ and Atlanta, GA.^19–21^ Both models compare outcomes between non-Hispanic Black and non-Hispanic White MSM. Modelling HIV among the substantially larger MSM population in LAC necessitates the inclusion of a third major group, Hispanic MSM, given the unique racial/ethnic composition of LAC.

We therefore developed a race/ethnicity-stratified microsimulation model for MSM that considers subgroup-specific partnership patterns, disease progression, patterns of PrEP use, and viral suppression outcomes from ART adherence patterns. We used a microsimulation to allow HIV disease and treatment dynamics (rates of transmission, diagnosis, treatment adherence, death, etc.) to vary by individual characteristics (race/ethnicity and age). Besides examining infection outcomes, we additionally calculated equality indices (Gini index, etc.) to evaluate the equality of outcomes across the examined policies. To our knowledge, this is the first publication using this type of analysis to examine PrEP allocation.

## METHODS

### Model Overview

We developed a discrete time microsimulation model to describe the transition of MSM in LAC between health and treatment states. We restricted the model to MSM, as this group alone accounted for 83% of new HIV diagnoses in 2019.^4^ Each health state is a collection of attributes that define an individual’s infection status and disease state (i.e., no infection, CD4 >= 500, 200 <= CD4 <= 499, CD4 <= 199), viral suppression (i.e. HIV-1 RNA < 200 copies/mL), PrEP usage (i.e., actively on a PrEP prescription), and diagnosis status (i.e., aware versus unaware if HIV positive). Transitions between health states was determined by annual transition probabilities drawn from empirical data, derived from prior literature, or determined via model calibration (see below), and may vary by age (15-100) and race/ethnicity (non-Hispanic Black, Hispanic, and non-Hispanic White).

We used data from 2011 to initialize our simulation and data from 2012-2016 for calibration. For model simplicity and due to data limitations, we did not include other racial/ethnic minority MSM as they comprise a very small portion of the PLWH in LAC: each racial/ethnic minority group constitutes less than 5% of PLWH in LAC (for a combined total of <10%). The UCLA and LAC Institutional Review Boards have approved this study for IRB Exempt status as all empirical data used were deidentified (IRB#19-000110).

The model used yearly cycles. Each year, men enter the model at age 15. Each year, individuals could acquire HIV, or if HIV-positive, progress through the stages of HIV infection. We did not consider immigration, emigration, or time of same-sex sexual debut. In addition to non-virally suppressed HIV prevalence, the risk of acquiring HIV depends on the individual and his sexual partners’ count, PrEP usage, demographic characteristics of age and race/ethnicity, and level of viral suppression in the community based on ART adherence. HIV-negative MSM may be prescribed PrEP, and PLWH may be diagnosed and enter viral suppression through ART treatment. Discontinuation and suboptimal adherence can occur for PrEP and virally suppressed ART users. Men can exit the model through death (either natural or AIDS-related)

Figure 1 depicts the health states and transitions for a single age- and racial/ethnic group (only one of the age and race/ethnicity combinations from the model is shown, for illustrative purposes). Individuals in the susceptible state are HIV-negative and not on PrEP. If infected, they can progress through HIV stages defined by CD4 count status. Those who are HIV-negative but seek PrEP may start PrEP, which reduces the likelihood of acquiring HIV. Each year, HIV-negative individuals can start PrEP and individuals on PrEP can stop. When someone acquires HIV, regardless of CD4 level, they will stop using PrEP when diagnosed and may become virally suppressed using ART. Patients who are virally suppressed from ART may be highly adherent through the year (95% likelihood), which results in no transmission of HIV. An individual’s HIV status, PrEP usage, viral suppression, and diagnosis status changes his probability of acquiring HIV, becoming diagnosed, and transmitting HIV. Appendix sections 1-4 provides additional details.

**Figure 1.**
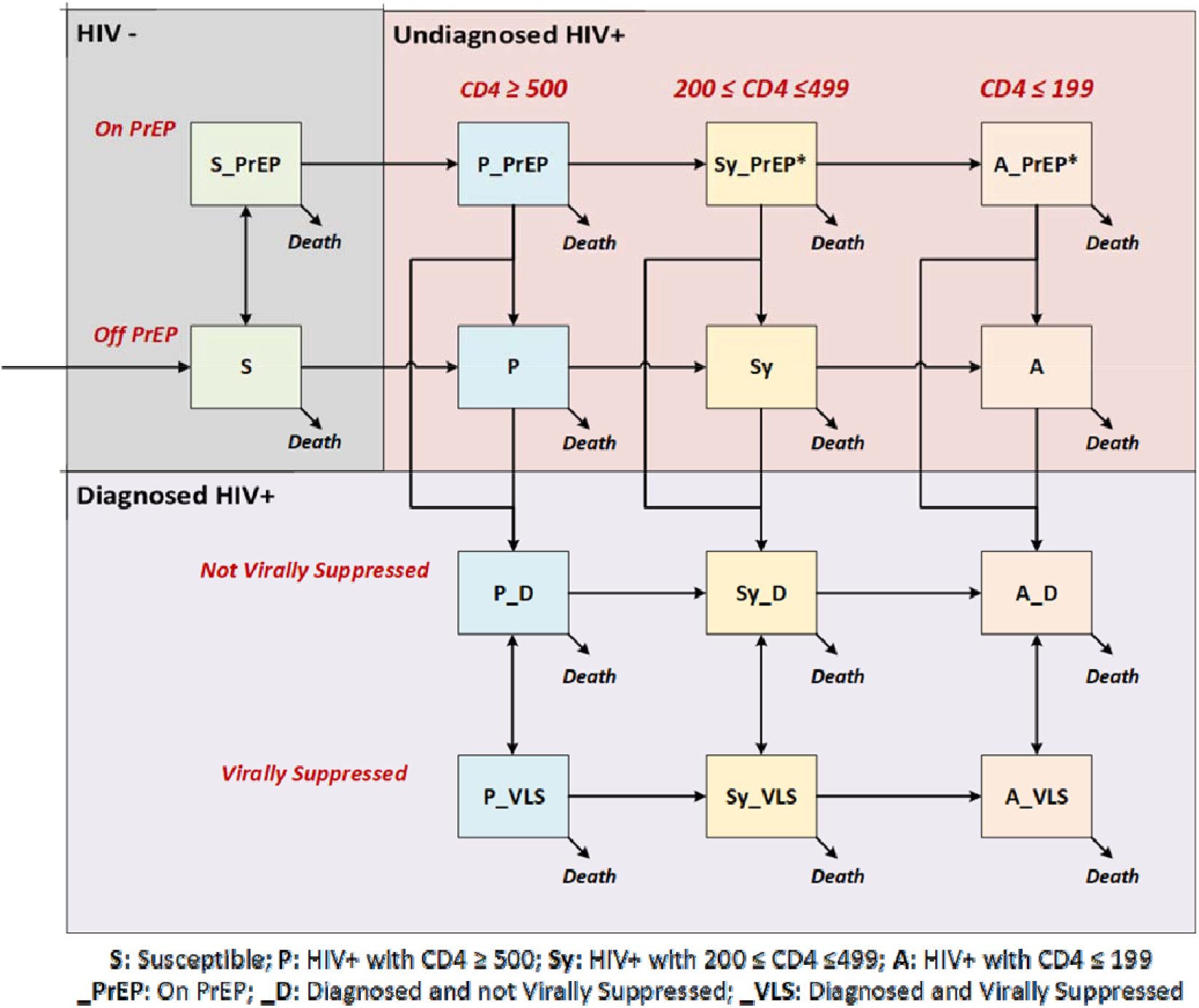
This simplified model schematic reflects disease and treatment progression for one age- and racial/ethnic-group (all combinations are modeled but omitted from the diagram for clarity). Arrows represent transitions that can occur within a particular age and racial/ethnic group. Individuals who have been diagnosed cannot be on PrEP. Sy_PrEP and A_PrEP health states have fewer than 5 individuals but are shown for completeness.

### Model Features and Inputs

We drew model inputs from empirical data and prior published work. Selected values are shown in Table 1 (complete list in Appendix section 2).

**Table 1:**
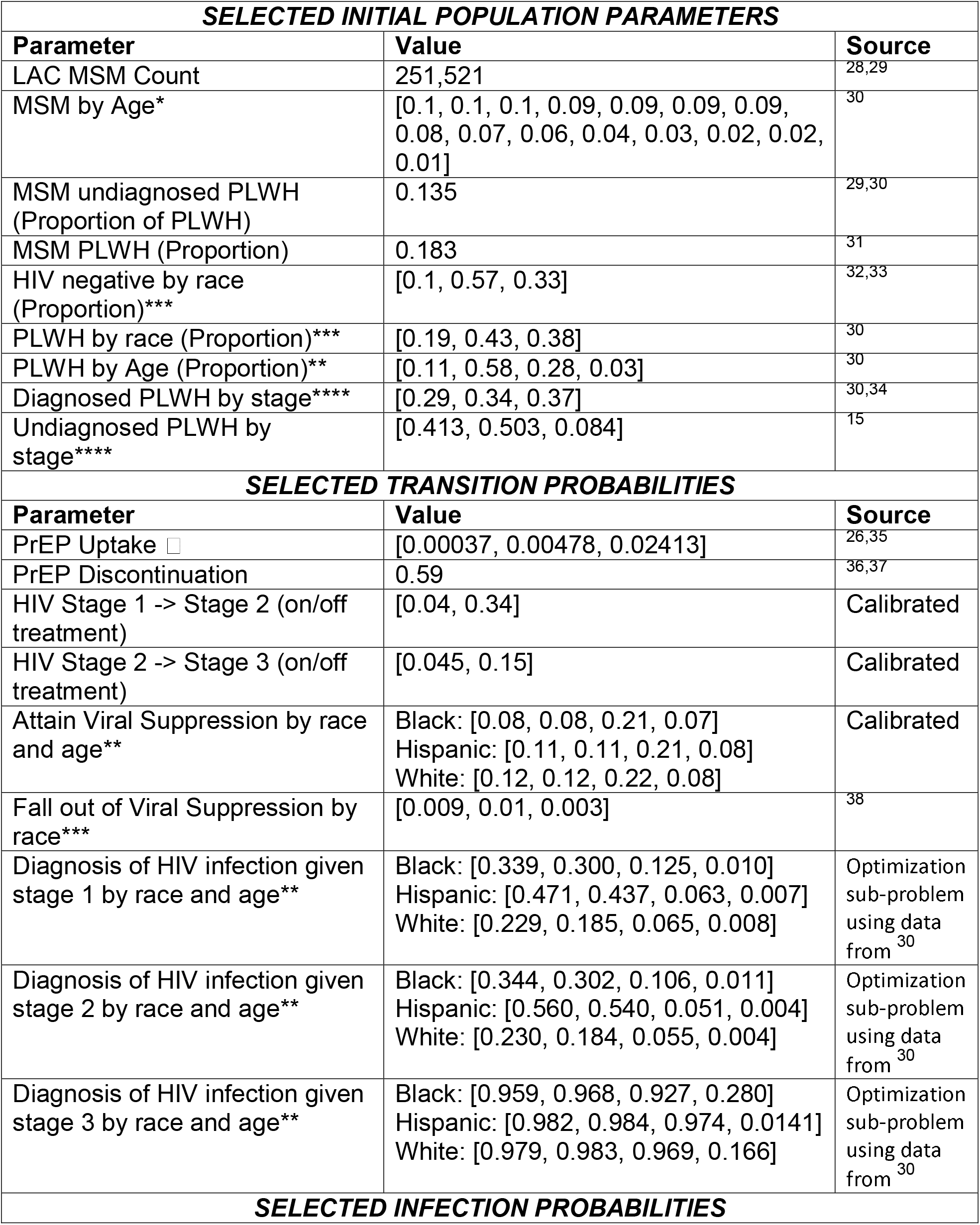
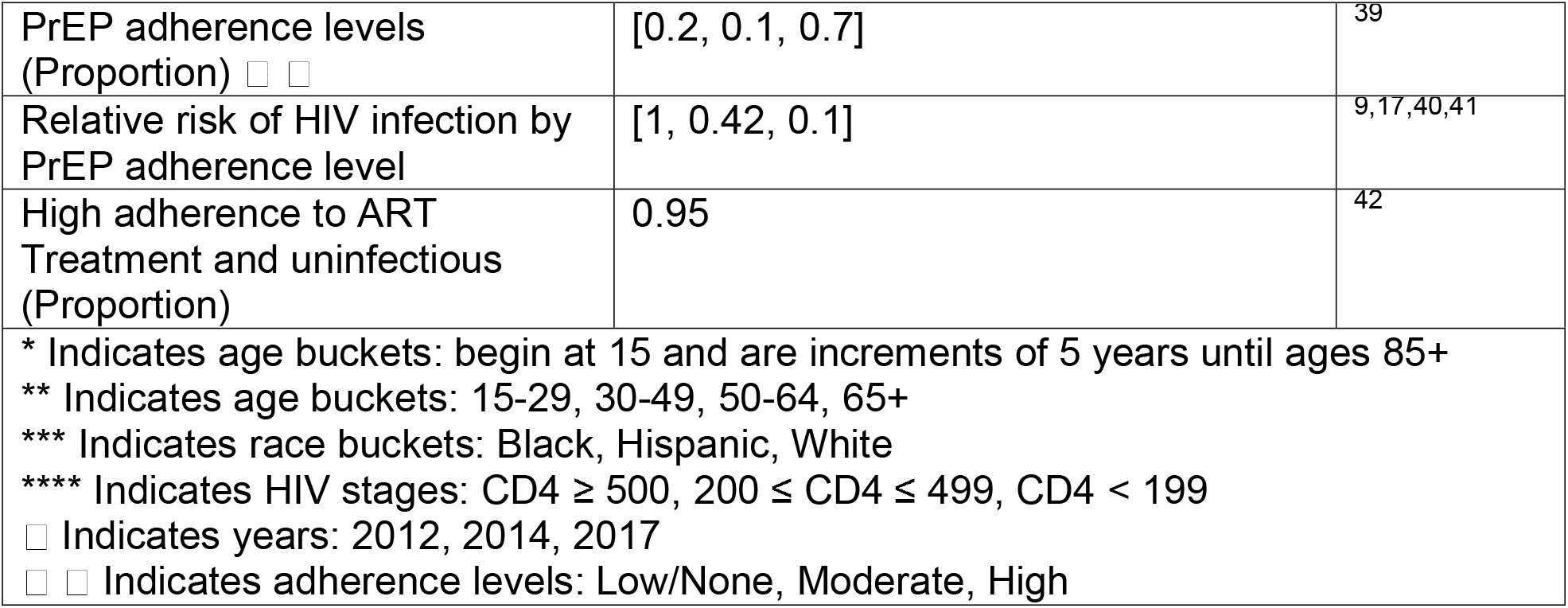
Selected Parameters (Complete list of parameters can be found in Appendix Sec. 2)

The annual probability of new HIV infection depends on an individual’s race/ethnicity and age, number of partnerships, PrEP status and adherence, and population characteristics (number of infectious PLWH and their race/ethnicity, age, and ART status and adherence) [see Appendix 2e]. These values were calculated in part from a partnership survey conducted by the LGBT Center of LA,^22^ shown in Figure 2.

**Figure 2:**
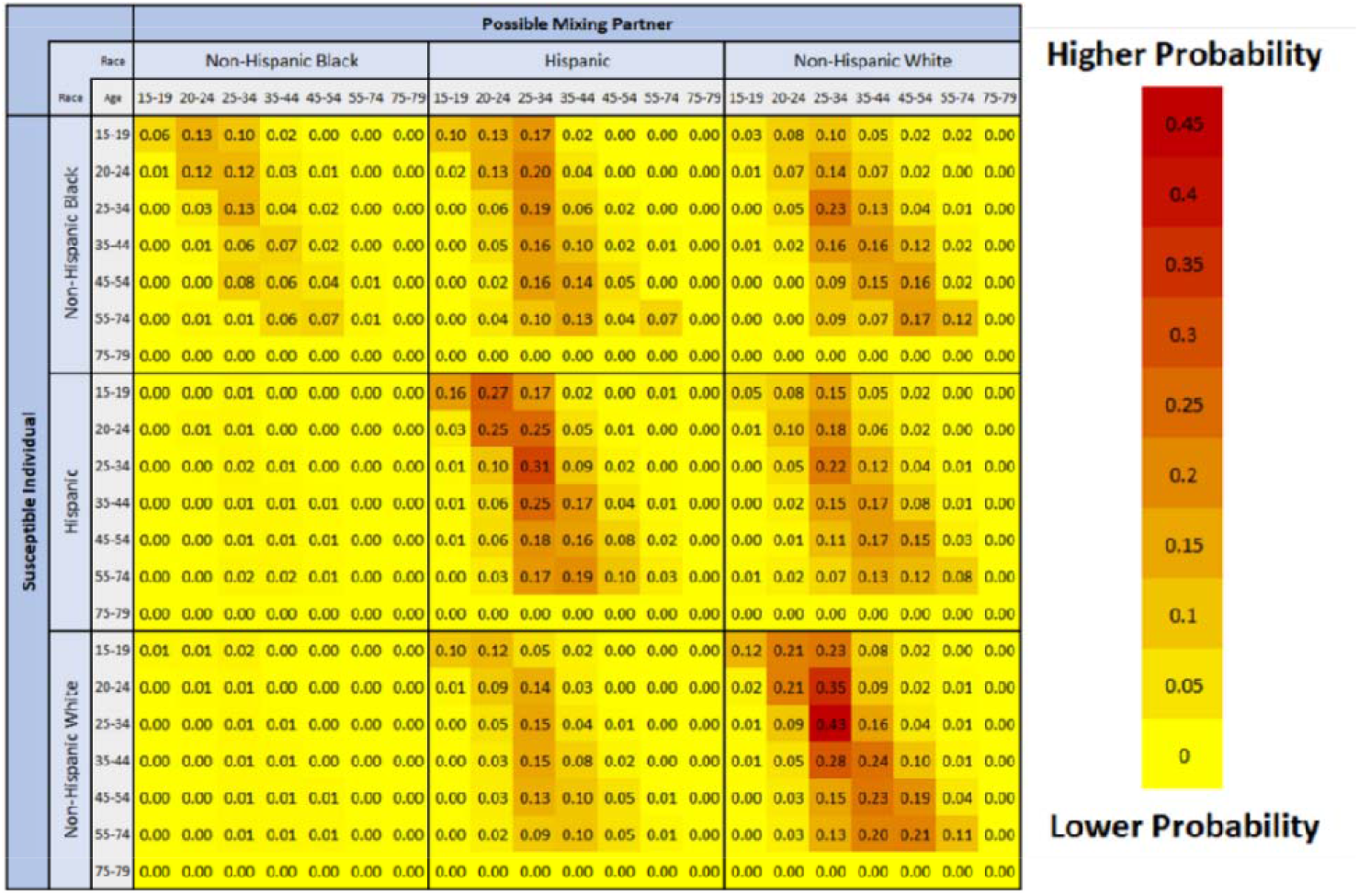
Partnership matrix. Rows represent the age and race/ethnicity of the susceptible individual, and columns the age and race/ethnicity of the possible partner.

This partnership matrix may help capture some of the complex factors that drive partnership mixing among MSM in LAC (e.g., preferences, neighborhood segregation, social/sexual racism). These factors explain, in part, some differences in HIV rates between racial/ethnic groups. Transmission patterns may greatly influence model forecasts, so accounting for these factors is critical. Since these race/ethnicity and age partnership patterns can be subject to bias, we performed sensitivity scenarios to determine model robustness.

### Model Calibration and Validation

Calibration was performed over the 2012-2016 period to determine values for uncertain parameters that we were unable to estimate directly. Calibrated values include calibration constants to account for relative risks, reaching viral suppression through ART by race/ethnicity and age, and disease progression while virally suppressed or not. We used a hierarchical calibration process with 35 calibration targets observed in the LAC Department of Public Health surveillance data (details in Appendix section 3). To validate the model and benchmark outcomes to local and national values, we compared 19 different model outputs to CDC data and published literature/reports of HIV prevalence, incidence, viral suppression, new diagnoses, HIV status awareness and PrEP for overall and race/ethnicity-specific values (see Appendix Section 4).

### Policy Scenarios

We simulated 18 strategies where an additional fixed number of PrEP initiation prescriptions (i.e., 3000, 6000, and 9000 coverage levels) are allocated annually across different racial/ethnic groups for 15 years (2021-2035). These PrEP prescriptions are given in addition to the status quo PrEP use, which we assumed was maintained over the simulated horizon. This was meant to proxy uneven PrEP uptake across groups, as may occur if the additional PrEP prescriptions are distributed by clinics or other resource-providing organizations that primarily service specific racial/ethnic groups (e.g., due to location or other factors); or if outreach encouraging PrEP uptake varied in effectiveness across different communities. For reference, under the baseline PrEP uptake (no intervention), approximately 4,500 individuals started PrEP in 2020. The additions of 3000, 6000, and 9000 prescriptions therefore increased the amount of PrEP prescribed by approximately 67%, 133%, and 200% respectively, relative to 2020.

We tested three allocation scenarios for distributing these additional PrEP prescriptions across three racial/ethnic groups of MSM: (1) Equal allocation (Equal quantity of PrEP for each group), (2) Count allocation (Proportional allocation based on the number of PLWH in each group), and (3) Rate allocation (proportional allocation by the new diagnosis rate in each racial/ethnic group). We also simulated scenarios where the additional PrEP is allocated to only one racial/ethnic group to better understand policy outcomes (detailed policy descriptions in Appendix Section 5).

### Model Outcomes

For each scenario, we identified the cumulative infections averted and new infection rates per 100,000 population in 2035 relative to a case where no intervention was implemented. We reported average values over 30 iterations per scenario, which was sufficient to generate small standard errors.

We also examined the equality in distribution of new infection rates in 2035 though the Gini index,^23,24^ a commonly used measure of equality. We provided other accepted equality indices (Atkinson and Kolm),^25^ with definitions, in Appendix Section 7. We additionally compared Gini index and incidence rates per 1,000 PrEP (e.g., the reductions are divided by 3 in the 3000 PrEP policies, by 6 in the 6000 policies, etc.) to assess whether tradeoffs in health and equity occur.

### Sensitivity Analyses

We performed sensitivity analyses on transmission patterns, as these values were inferred from a non-representative sample. We used two alternative partnership mixing scenarios: (1) Assortative mixing: individuals only have partners of the same racial/ethnic group, with no age preferences; (2) Uniform mixing: individuals have equal likelihood for a partner of any other age and racial/ethnic group. See Appendix section 7.

## RESULTS

### Calibration and Validation Results

The root mean squared error (RMSE) of average percent error over aggregate calibration targets were all below 10%, indicating that the model shows good calibration for most targets by race/ethnicity, age, and HIV stage (Appendix Table 8). See Appendix Section 3 for details.

We found that our model performs within 10% of the values reported in the literature for undiagnosed PLWH, viral suppression, new diagnoses, total PLWH, incidence rate, and PrEP Coverage.^26–28^ We also found that our model performs well when comparing relative incidence rates, relative diagnosis rates, HIV status awareness, and viral suppression across racial/ethnic groups to LA County trends (see Appendix Table 10 and Appendix Section 4).^4^

### Policy Outcomes

Enrolling 9000 additional PrEP users annually among Black MSM averted the most cumulative infections from 2021-2035. Figure 3 depicts the number of cumulative infections averted relative to no intervention. As anticipated, larger increases in PrEP coverage resulted in greater overall benefit under all allocation schemes. At high allocation levels among Black MSM, there were “spillover” effects to Hispanic MSM as secondary infections among Hispanic MSM are averted. PrEP allocation to Hispanic or White MSM groups did not show the same spillover effects. See Appendix Table 13 for values and standard errors.

**Figure 3:**
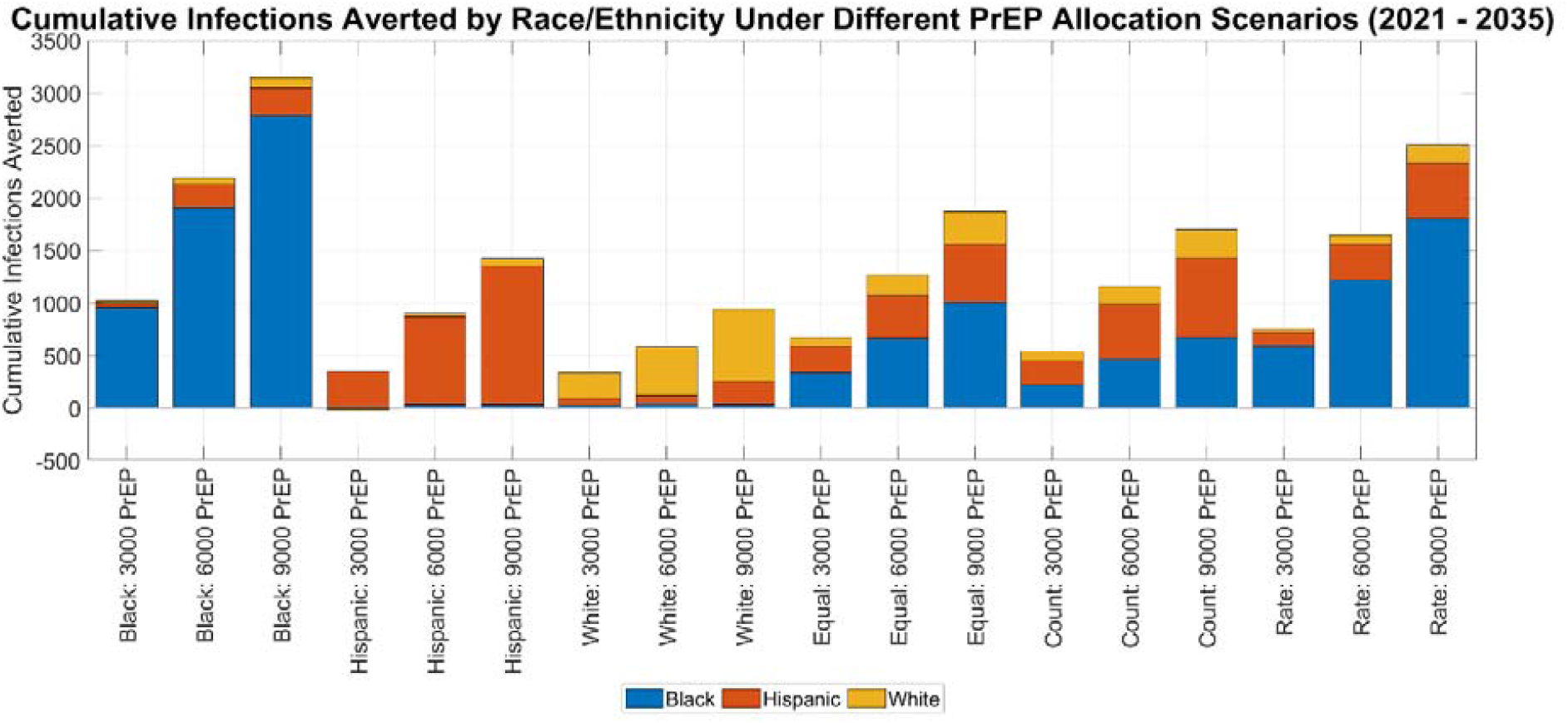
Cumulative infections averted for all simulated policies. Results indicate where the benefit was observed in the population by race/ethnicity. Note that each policy is defined by the allocation scheme and additional annual PrEP coverage.

Within the distributed allocations (Equal, Count, and Rate), the Rate policy – allocation proportional to new HIV diagnoses in that group – distributed most of the PrEP to Black MSM, while the Count policy – allocation proportional to size of PLWH population in that group – distributed the least to Black MSM. At all three coverage levels, the Rate policy averted more cumulative infections than the other policies.

### 2035 Outcomes

Unsurprisingly, larger PrEP intervention policies garnered larger reductions in 2035 incidence rates (see Figure 4a). The policy targeting Black MSM with 9000 PrEP reduced overall incidence the most, to approximately 550 new cases per 100,000 susceptible MSM. However, all policies reduced incidence rates from the No Intervention case.

**Figure 4:**
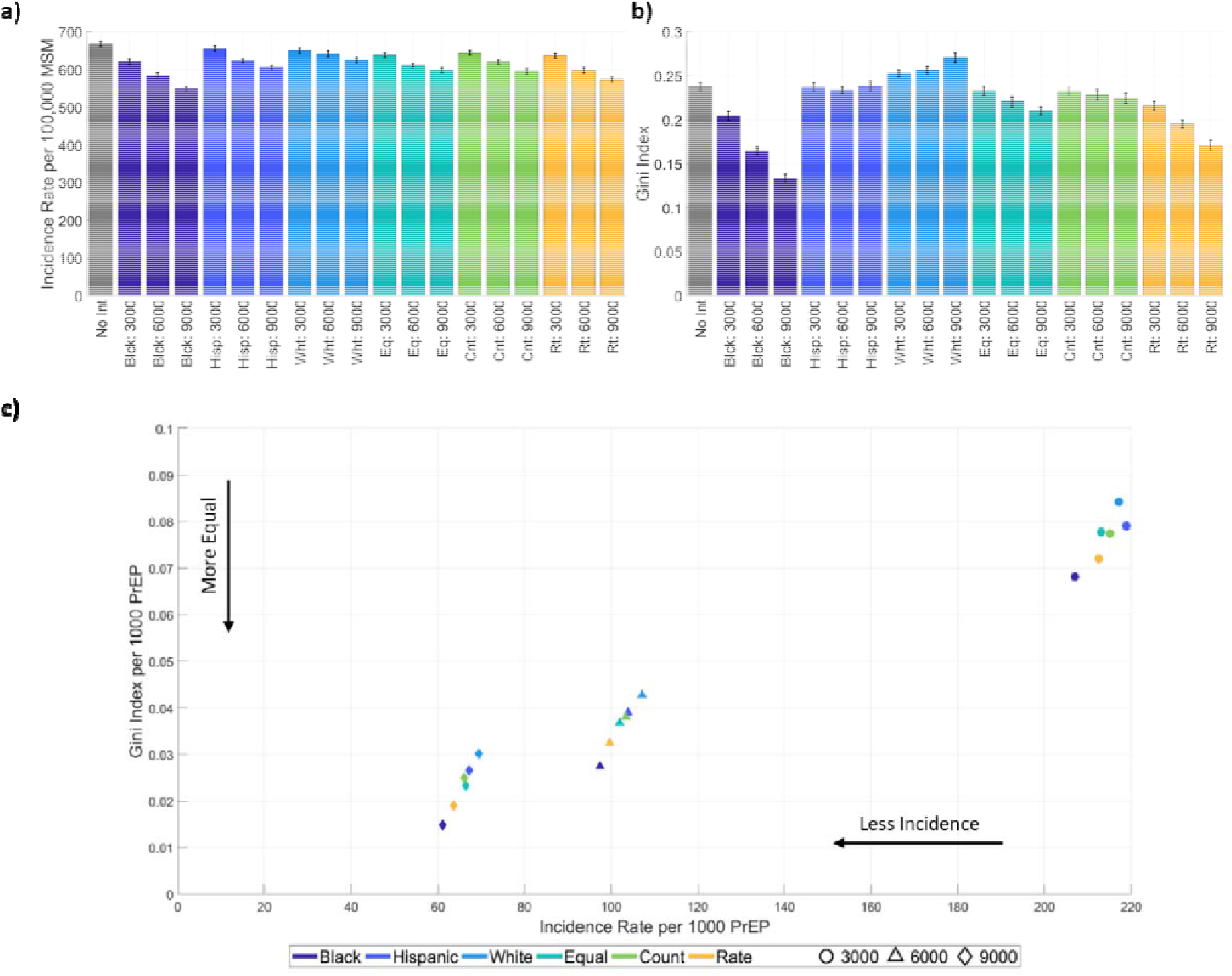
Panel (a) shows the Gini index for incidence rates in 2035 for all allocation and PrEP level combinations (see Appendix section 5 for definition). Panel (b) shows incidence rates in 2035 per 100,000 susceptible MSM. Panel (c) shows the outcomes in equality and effectiveness for the simulated policy per 1000 additional annual PrEP for comparability between intervention levels. Abbreviations: No Int = No Intervention, Blck = Black only policy, Hisp = Hispanic only policy, Eq = Equal policy (distributed equally to all racial/ethnic groups), Cnt = Count policy (distributed proportionally to the total PLWH in each racial/ethnic group), and Rt = Rate policy (distributed proportionally to the new diagnosis rates in each racial/ethnic group).

Targeting only Black MSM also generated a more equal distribution of incidence rates across the population. The Gini, Atkinson, and Kolm indices showed similar trends, so we only show the Gini index outcomes in Figure 4b (see Appendix Section 7 for the others). Using all equality metrics, we found that the policy targeting Black MSM with 9000 additional PrEP resulted in the most equal outcomes of the policies we evaluated in 2035. By contrast, the other single-race policies, targeting Hispanic or White MSM only, led to roughly equal or higher Gini Indices compared to having no intervention.

All policies besides targeting White MSM increased both effectiveness and equality compared to no intervention. Allocating 9000 additional PrEP resulted in lower Gini index and incidence rate, per unit of PrEP, than the 6000 and 3000 interventions. Within each intervention level (3000, 6000, or 9000), targeting Black MSM always garnered the lowest levels of Gini index and incidence rate (see Figure 4c).

## Results of Sensitivity Scenarios

The results were sensitive to partnership mixing assumptions, and the numbers of infections averted varied widely. However, we found that the trends in cumulative infections averted were similar to the empirical mixing results, with policies that prioritize PrEP allocation to Black MSM averting the most cases (see Figure 5). The benefits of the intervention were more evenly distributed across groups under uniform mixing and less so under assortative mixing. As expected, assortative mixing generated no spillover effects while uniform mixing showed substantial spillover effects. Differences in averted cases across mixing scenarios were driven by differences in incidence rates under the No Intervention policy. Incidence rates were 1.5 to 4 times higher for non-Hispanic Black MSM in the assortative mixing scenario compared to the empirical mixing scenario; similarly, incidence rates were 1.25 to 2 times higher for Hispanic MSM under uniform mixing. Additionally, under uniform mixing, we did not find a decline in incidence rate by 2035, as we saw under empirical mixing (see Appendix Section 7).

**Figure 5:**
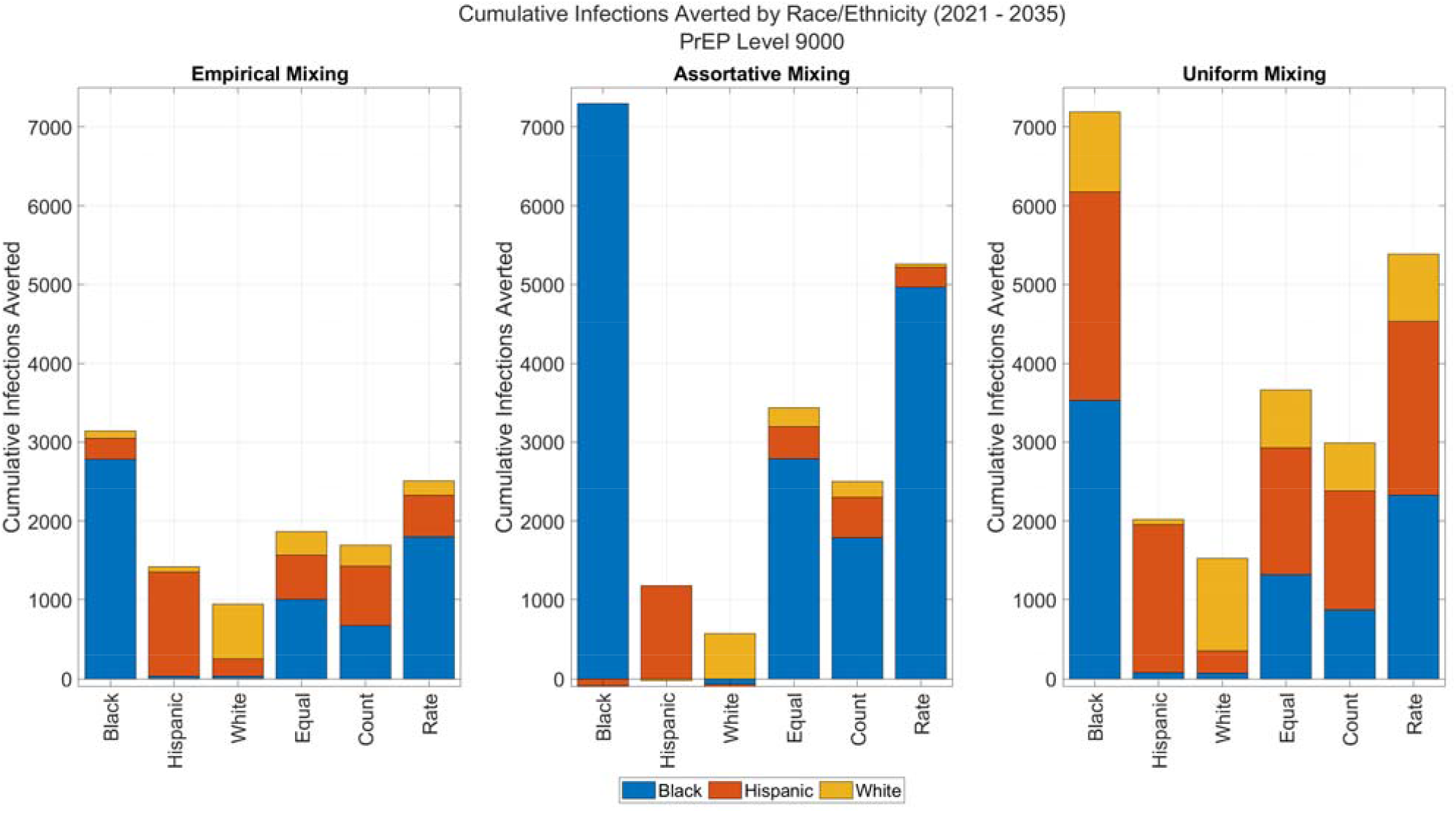
Cumulative infections averted for all allocation scenarios at the 9000 level under three different mixing patterns. Results indicate where the benefit was observed in the population by race/ethnicity. Empirical mixing is based on data collected from an LA LGBT center clinic. Assortative mixing assumes partnerships only exist within the same race/ethnicity. Uniform mixing assumes partnership preferences are equal across all age and race/ethnicities.

Although these mixing scenarios result in differences in the Gini index at all policy levels, the trends remained consistent to those seen under empirical mixing. The Black and Rate policies level reduced disparities substantially (Gini index reduction of 39%-53% and 28%-33%, respectively, at the 9000 level), while the Equal and Count policies resulted in small benefits (reduction of 11%-13% and 6%-9%). The Hispanic and White policies consistently maintained or increased disparities.

## DISCUSSION

We developed a race/ethnicity- and age-stratified microsimulation model to assess the potential epidemiological and equity impacts of 18 alternative PrEP allocation strategies among MSM in LAC. We found that parity in incidence rates across racial/ethnic groups will not be attained by 2035 if no additional policies are implemented. While efforts in the past decade have reduced disparities, there is much work yet to be done. None of the interventions examined here were able to achieve parity in incidence rates by 2035, despite doubling PrEP from 2020 levels in the largest interventions.

Our results suggest that a policy targeting Black MSM for PrEP can generate the most cumulative reduction in new cases over the next 15 years – and that doing so would also reduce the gap between incidence rates between Black MSM and MSM of other racial/ethnic groups. Increasing PrEP coverage by 9000 additional prescriptions annually to Black MSM would avert approximately 3140 HIV infections by the year 2035, with incidence rates of approximately 720, 650, and 350 per 100,000 among Black, Hispanic, and White MSM, respectively, in 2035. This represents a much smaller disparity gap than the projected incidence with no policy intervention (1940, 680, and 380 per 100,000 MSM in these groups, respectively). More modest PrEP scale-up, while generating lower cumulative benefits, shows similar trends in reducing disparities when focused on Black MSM. These results are reinforced by examining equality measures such as the Gini index, which estimates that targeting Black MSM for PrEP could reduce the Gini Index to 0.13 in 2035 from 0.24 with no intervention, a 46% reduction.

Of the policies that do not target only a single race/ethnicity, the Rate policy, which allocates PrEP by diagnosis rate in 2021, averts the most HIV infections and decreases disparities the most. At the 9000 PrEP level, it results in approximately 2500 cumulative cases averted and final incidence rates of 1090, 630, and 350 per 100,000 in 2035 among Black, Hispanic, and White MSM, respectively.

We also examined the relationship between equality and effectiveness of PrEP policies. While all policies were dominated by those targeting Black MSM for PrEP, we found that all policies besides targeting White MSM improved both overall incidence rate and the Gini index, suggesting that a tradeoff between equality in outcomes and effectiveness is generally not a concern in the interventions we evaluated. This is likely because policies that target by race/ethnicity reduce incidence rates the most in groups that bear disproportionate disease burdens.

These results are dependent on our model inputs and assumptions, as demonstrated by our sensitivity analysis around mixing patterns. Changes in mixing patterns greatly change model projections of incidence and disparities. The likelihood of averting cases in other racial/ethnic groups when one group is targeted depends strongly on partnership patterns. However, regardless of mixing patterns, we found that targeting Black MSM for additional PrEP prescriptions continued to result in the largest cumulative infections averted and the lowest disparities between groups in 2035, suggesting that this result is robust to even extreme changes in model assumptions.

### Limitations

This study has several limitations. First, we drew from multiple data sources, including county-specific surveillance data for MSM, published literature and models, CDC reports, and others at various levels of stratification by age, race/ethnicity, and treatment. Use of disparate data sources can result in possible data discrepancies and there may be uncertainty in the surveillance data as reporting practices change over time. Second, due to lack of data on multiple characteristics simultaneously, we used a quadratic programming approach to infer the joint distributions, assumptions of independence, or assumed that the parameter did not vary by demographic characteristics. While this approach may not perfectly accurately capture all demographically correlated trends, it provides our best estimate given available data. Third, we used non-representative survey data on partnerships to define the partnership matrix, as this was the best data available.^22^ We mitigated this limitation by conducting sensitivity analyses on partnership patterns, which revealed that cumulative infections averted can vary substantially under different partnership mixing. Fourth, we did not consider mental health status, substance use, housing status, and other risk factors (beyond age and race/ethnicity) that have been shown to influence HIV risk, ART adherence, and PrEP uptake. Incorporating these additional factors would require substantially more data, much of which may not be available. Additionally, the influence of these factors may be indirectly captured in the model, insofar as they are correlated with age and race/ethnicity.

## Conclusions

Despite these limitations, our analysis provides important insights into the relationship between effectiveness and disparity reduction across a variety of PrEP policies. We quantified equality of outcomes using widely accepted indices, providing comparable metrics for evaluating the relative equality benefits of the policies evaluated. This allowed us to examine the relationship between equality and overall incidence, which showed that most policies we examined were able to reduce inequality and incidence simultaneously. In addition, we found that targeting Black MSM dominated other policies at all intervention levels we considered.

We improved upon existing models by disaggregating by age and race/ethnicity and incorporating empirical data on partnership mixing patterns. While imperfect, this approach may capture partnership mixing patterns that are influenced by a variety of social factors, including segregation and racism. This treatment of mixing within the model therefore represents a substantial advance in how sexual partnerships are represented. These partnership dynamics also allow for a more nuanced understanding of the downstream effects of averted HIV infections through PrEP uptake. To the best of our knowledge, this microsimulation model is also the first to reflect LAC demographics, with stratifications for age and race/ethnicity.

Models like this one can enable policymakers to assess tradeoffs between the dual goals of reducing overall HIV burden and reducing inequalities. Simultaneous achievement of these aims is integral towards achieving EHE objectives at the local level. However, health gains and inequality reduction objectives must be balanced against the costs of policies and programs. These may include, for example, differential costs related to outreach to different population subgroups and distribution of resources across the portfolio of HIV prevention and treatment strategies. Recommended strategies may differ after consideration of these tradeoffs. The insights from this analysis will be useful in informing the discussion around strategies to reduce racial/ethnic disparities in HIV/AIDS burden, prevention, and care.

## Data Availability

Model parameters are provided in the attached appendix

## Notes

**Financial Support:** This work was supported by the California HIV/AIDS Research Program (RP15-LA-007) and the UCLA Center for HIV Identification, Prevention and Treatment Services (P30 MH058107), CHIPTS, and the UCLA Center for AIDS Research (CFAR), UCLA AIDS Institute and USC Keck School of Medicine Seed Grant for Cross-Campus Collaboration. The funding sources had no role in the study design, data analysis and interpretation, decision to publish, or preparation of the manuscript.

### Competing Interest Statement

The authors have declared no competing interest.

### Funding Statement

This work was supported by the California HIV/AIDS Research Program (RP15-LA-007) and the UCLA Center for HIV Identification, Prevention and Treatment Services (P30 MH058107), CHIPTS, and the UCLA Center for AIDS Research (CFAR), UCLA AIDS Institute and USC Keck School of Medicine Seed Grant for Cross-Campus Collaboration. The funding sources had no role in the study design, data analysis and interpretation, decision to publish, or preparation of the manuscript.

### Author Declarations

UCLA and LAC Institutional Review Boards have determined this study to be exempt (IRB#19-000110).

