## Supplemental Appendix for "Impact and Equality of Expanding Pre-Exposure Prophylaxis (PrEP) for Men who have Sex with Men in Los Angeles County"

#### 1. Microsimulation Structure

The microsimulation was developed to model HIV progression and transmission of HIV among men who have sex with men (MSM) in Los Angeles County. In the simulation, individuals transition between health and treatment states to estimate HIV prevalence, incidence, and treatment outcomes over a 15-year time horizon (2021-2035). It is Markovian with a one-year cycle time. To ensure that the model reflects reality, it starts in 2011 and data from 2012-2016 are used to calibrate the model. All transition probabilities are constant over the simulation time horizon except for PrEP uptake, which is time variant. PrEP uptake is made time variant because of how significantly PrEP usage trends have changed since it was first approved in 2012.

The simulated population reflects the characteristics of the Los Angles (LA) County MSM population in both race/ethnicity (normalized non-Hispanic Black, Hispanic, non-Hispanic White) and age (15-100). We do not consider the migration of MSM into or out of Los Angeles County or discovery of sexual orientation towards population growth; individuals only enter the simulation via aging. Exiting the microsimulation only occurs via death. All individuals in the population can be described by the attributes specified in Table 1.

*Table 1: Simulation individual attributes*

| **Attribute** | **Characteristics** |
|---|---|
| Age | • Defined on entrance to the model<br>• Increases by 1 every year until death<br>• Death occurs after aging if died in a given year |
| Race/Ethnicity | • Defined on entrance into model<br>• Never changes |
| HIV Status | • 4 possible statuses<br>• HIV negative, CD4= > 500, 200 <= CD4 <= 499, CD4 <=200<br>• Progression is independent of race and age |
| PrEP Usage | • PrEP usage can have variable levels of adherence<br>• Can only be on PrEP if not diagnosed<br>• Can only start PrEP if susceptible<br>• Can switch between on and off PrEP if susceptible<br>• Cannot be on PrEP and treatment |
| Diagnosed | • Defined as someone who has HIV and is aware of it, NOT as someone who has been tested<br>• HIV negative individuals will never be categorized as diagnosed<br>• Cannot transition out of being diagnosed with HIV |
| Viral Suppression | • Can only be virally suppressed if diagnosed<br>• Can switch on and off viral suppression<br>• If viral suppression, cannot be on PrEP<br>• Viral suppression be attained at any stage except susceptible<br>• Viral suppression measures are based by end of year |
| Alive | • Can die either naturally or by AIDS<br>• All other attributes remain constant after death |

In Figure 1 of the manuscript, we present a model diagram showing progression of individuals through health states in the microsimulation. Boxes represent health states while arrows represent transitions between health states. Note that transitions may vary based on age and race. It is unlikely an individual progresses all the way to A_PrEP as they are likely to be diagnosed prior to this point.

*Table 2: Model Abbreviations*

| **Health State** | **Abbreviation** |
|---|---|
| HIV negative and not on PrEP | S |
| HIV negative and on PrEP | S_PrEP |
| HIV positive (CD4 >= 500), undiagnosed, and not on PrEP | P |
| HIV positive (CD4 >= 500), undiagnosed, and on PrEP | P_PrEP |
| HIV positive (CD4 >= 500), diagnosed, and not Virally Suppressed | P_D |
| HIV positive (CD4 >= 500), diagnosed, and Virally Suppressed | P_VLS |
| HIV positive (200 <= CD4 <= 499), undiagnosed, and not on PrEP | Sy |
| HIV positive (200 <= CD4 <= 499), undiagnosed, and on PrEP | Sy_PrEP |
| HIV positive (200 <= CD4 <= 499), diagnosed, and not Virally Suppressed | Sy_D |
| HIV positive (200 <= CD4 <= 499), diagnosed, and Virally Suppressed | Sy_VLS |
| HIV positive (CD4 <= 200), undiagnosed, and not on PrEP | A |
| HIV positive (CD4 >= 500), undiagnosed, and on PrEP | A_PrEP |
| HIV positive (CD4 >= 500), diagnosed, and not Virally Suppressed | A_D |
| HIV positive (CD4 >= 500), diagnosed, and Virally Suppressed | A_VLS |

The following transitions exist in the microsimulation and occur in the defined order : (1) New diagnoses, (2) acquiring infection, (3) HIV status progression, (4) PrEP adoption/cessation, (5) Adoption/cessation of treatment at levels to reach viral suppression, (7) intervention, (6) aging, (7) death. The order in which these transitions are performed is significant as the changes caused by one transition impact the state of the individual for the next transition. For example, PrEP and treatment adoption/cessation transitions occur sequentially after new diagnosis, acquiring infection transitions, and HIV status progression because these parameters impact the probability of PrEP uptake and starting treatment. An individual would not start PrEP if they were HIV positive and an individual would not start treatment unless they had been diagnosed.

### 2. Model Parameters

#### 2a. Initial Population

Because HIV trends are not in steady state, a burn-in procedure for the simulation would not be an appropriate method for determining the characteristics of the initial population. We find that data on characteristics for the MSM population are scarce. When MSM specific data is not available, we assume general population trends to our MSM community. Additionally, many of the metrics presented in literature or reports are not given by race and age, as needed for the simulation, so we either assume independence between these parameters or develop optimization subproblems to identify a feasible joint distribution to apply. In general, we use data specific to LA county wherever possible. When not available, we use proportions at the state or national level. Similarly, we aim to use MSM-specific parameters when possible but use male-specific or general population characteristics when data is not provided for MSM specifically. In Table 3, we present the initial population parameters.

*Table 3: Initial population parameters*

| **Parameter** | **Value** | **Range** | **Source** |
|---|---|---|---|
| LA County MSM Count | 251521 | [234,636 – 300,628] | [1,2] |
| LA County Proportion of Male Population by Age* | [0.1, 0.1, 0.1, 0.09, 0.09, 0.09, 0.09, 0.08, 0.07, 0.06, 0.04, 0.03, 0.02, 0.02, 0.01] | | Calculated from data provided by LACDPH [3] |
| LA County Proportion of Population by race (Proportion)** | [0.1, 0.57, 0.33] | | [4,5] |
| MSM Diagnosed PLWH (Proportion) | 0.158 | | Calculated from [1,3] |
| MSM undiagnosed PLWH (Proportion of PLWH) | 0.135 | [0.108, 0.136] | Table 2 of [6] |
| MSM PLWH (Proportion) | 0.183 | [0.07, 0.371] | Calculated from [7] |
| PrEP Coverage (2011) | 0 | | [8] |
| Diagnosed PLWH Virally suppressed given race (Proportion)*** | [0.44, 0.56, 0.59] | | [3] |
| Diagnosed PLWH Virally suppressed given age (Proportion)** | [0.40, 0.54, 0.62, 0.63] | | [3] |
| PLWH by race (Proportion)*** | [0.19, 0.43, 0.38] | | [3] |
| PLWH by Age (Proportion)** | [0.11, 0.58, 0.28, 0.03] | | [3] |
| Diagnosed PLWH by stage**** | [0.29, 0.34, 0.37] | | [3,9] |
| Undiagnosed PLWH by stage**** | [0.413, 0.503, 0.084] | | [10] |

** Age breakdown stratifications begin at age 15 and are increments of 5 years until ages 85+*

*** Age breakdown for diagnosed population are 15-29, 30-49, 50-64, and 65+*

**** Race (Race/ethnicity) stratification considers non-Hispanic Black, Hispanic, and non-Hispanic White/Other*
**** Stages are based on the following CD4 levels: CD4 >= 500, 200 <= CD4 <= 499, CD4 <= 200*

The population at the beginning of the simulation (end of year 2011) consists of 251,521 MSM individuals based on an estimate reported by Grey using the American Community Survey , 2009-2013[2]. We assume that individuals are aged 15-100 years old. Hall's study reports approximately 13.5% of PLWH in the United States, 2008-2012, are undiagnosed[11]. Using these values and LA county surveillance data for diagnosed HIV cases in 2011, we estimate approximately 18% of the overall LA County MSM population was PLWH[3]. Demographic characteristics in the initial population (10% Black, 57% Hispanic, 33% White) follow values reported by the Census Bureau (2019)[4]. We formulate a quadratic programming optimization subproblem utilizing LA County Department of Public Health surveillance data on diagnosed HIV cases to determine the number of individuals in each age and race/ethnicity subgroup[3]. The proportion of undiagnosed individuals initialized to each HIV stage are taken from Khurana's national level HIV model[10]. The proportion of diagnosed individuals in each stage is estimated using county surveillance data[3].

Data on age breakdowns by race/ethnicity for MSM in Los Angeles County were not available. Therefore, for the susceptible and undiagnosed populations, we assume independence between the proportions identified for each stratification (age, race/ethnicity, and HIV stage). We then apply the joint proportion relevant for each subgroup to the overall population size to determine the count of individuals in all HIV negative and undiagnosed compartments.

For the diagnosed population, we have data on the number of individuals diagnosed with HIV by age and race for 2011, our initialization year, and the number of virally suppressed individuals by age and race. These values are provided by the LA County Department of Public Health from surveillance data and are specifically for MSM. Using these four values, we minimize the weighted sum of squared errors between imputed and empirical values to infer the breakdown of the diagnosed MSM population by race, age, and treatment. Using MATLAB CVX, we find a solution that yields an objective value that is approximately zero. We ensure that no compartments are empty.

#### 2b. Population Growth

Population growth in the microsimulation accounts for new entrants by aging. We assume that any population growth that would occur by immigration of MSM to LA county after age 15 or time of same-sex sexual debut is sufficiently low or offset evenly by MSM leaving LA county. This is consistent with other HIV modelling efforts with age stratifications[10]. Because we want to maintain the population of 15-year-olds as the population ages, the number of new entrants is equivalent to the approximate proportion of the population that is 15 in our initial population, 1.9%. Thus, for all years simulated, 1.9% of the prior year's population was added to the current year's population as 14-year-olds. These individuals enter the simulation prior to any simulated transitions for that year and are classified as 15-year-olds in the end of year metrics. All new entrants are considered HIV negative and none of them are on PrEP. The race/ethnicity proportional breakdown for new entrants align with the race/ethnicity breakdown used for HIV negative individuals in the initial population.

*Table 4: Population growth parameters*

| Parameter | Value | Source |
|---|---|---|
| New entrants (Proportion) | 0.019 | Calculated |
| New Entrants by race *(Proportion) | [0.1, 0.57, 0.33] | [4,5] |

** Race (Race/ethnicity) stratification considers non-Hispanic Black, Hispanic, and non-Hispanic White*

#### 2c. Transition Probabilities

Transition probabilities define the annual probability that an individual moves from one health/treatment state to another. These probabilities can be specific to a particular population subgroup, if the data was available (e.g., different probabilities for diagnosis, treatment, and PrEP use between non-Hispanics and Hispanics, etc.). This is one way in which the model can capture differences in behaviors that may exist between race/ethnicity and age groups. We assume that all age and race/ethnicity subgroups have the same likelihood of disease progression through the three HIV stages (with or without viral suppression) and the same likelihood of PrEP uptake and discontinuation. However, while PrEP uptake is the same across all subgroups, the likelihood of being prescribed PrEP changes from 2014 to 2017 to reflect the increase in PrEP adoption over time reflected in prior studies[12]. Natural death and the probability of dying of AIDS (virally suppressed or not) vary by age but are not race/ethnicity specific. By contrast, dropping from viral suppression is race/ethnicity specific, but not age. Other parameters such as reaching viral suppression and diagnosis probability at each stage (based on CD4 level) are both age and race/ethnicity specific. We are unable to make all transitions race/ethnicity and age specific due to limitations in existing data. While some transition probabilities are found through calibration (see section titled "calibration"), the others reflect values from prior literature and reports or derived from data provided by the CDC or LA County Department of Public Health. Table 4 outlines the transition probabilities used in the model.

*Table 5: Transition probabilities*

| Parameter | Value | Source |
|---|---|---|
| PrEP Uptake* | [0.00037, 0.00478, 0.02413] | [12,13] |
| PrEP Discontinuation | 0.59 | [14,15] |
| HIV Stage 1 -> Stage 2 (on/off treatment) | [0.04, 0.34] | Calibrated |
| HIV Stage 2 -> Stage 3 (on/off treatment) | [0.045, 0.15] | Calibrated |
| Attain Viral Suppression by race and age** | Black: [0.08, 0.08, 0.21, 0.07]<br>Hispanic: [0.11, 0.11, 0.21, 0.08]<br>White: [0.12, 0.12, 0.22, 0.08] | Calibrated |
| Fall out of Viral Suppression by race*** | [0.009, 0.01, 0.003] | Calculated from table 1 and 2 in [16] |
| Diagnosis of HIV infection given stage 1 by race and age** | Black: [0.339, 0.300, 0.125, 0.010]<br>Hispanic: [0.471, 0.437, 0.063, 0.007]<br>White: [0.229, 0.185, 0.065, 0.008] | Optimization sub-problem using data from [3] |
| Diagnosis of HIV infection given stage 2 by race and age** | Black: [0.344, 0.302, 0.106, 0.011]<br>Hispanic: [0.560, 0.540, 0.051, 0.004]<br>White: [0.230, 0.184, 0.055, 0.004] | Optimization sub-problem using data from [3] |

| | | |
|---|---|---|
| Diagnosis of HIV infection given stage 3 by race and age** | Black: [0.959, 0.968, 0.927, 0.280]<br>Hispanic: [0.982, 0.984, 0.974, 0.0141]<br>White: [0.979, 0.983, 0.969, 0.166] | Optimization sub-problem using data from [3] |

** PrEP uptake probability changes in 2012, 2014, and 2017*

*** Age categories are 15-29, 30-49, 50-64, 65+*

**** Race (Race/ethnicity) stratification considers non-Hispanic Black, Hispanic, and non-Hispanic White*

We use a quadratic programming minimization problem to identify the probability of an undiagnosed HIV positive individual becoming diagnosed, based on his race, age, and HIV stage. From the LA county surveillance data, we have counts for new diagnosis in 2012 based on race, age, and stage (independently).[3] From our initial population estimates, we also have estimated counts for the end of 2011 undiagnosed HIV positive individuals by HIV stage, race, and age. Age buckets are defined as 15-29, 30-49, 50-64, and 65+. The objective function used in the minimization problem is a sum of weighted squared errors. Similar to the diagnosed population count optimization problem described above, we solve the optimization problem using CVX in MATLAB and find a solution that yields an objective value that is approximately zero. We recognize the input data may contain measurement error. We ensure that no HIV stage, race, and age group have a probability of zero to be diagnosed.

#### 2d. Death Probabilities

Death probabilities in the simulation are age specific and derived from CDC data. All individuals who have not progressed to the stage of AIDS have an annual mortality probability based on age according to the 2016 CDC life table for males.[17] For those with AIDS, a life table is derived from the CDC mortality data for 2016.[17] We assume that this life table applies to those on treatment. Treatment has been reported to reduce HIV mortality by 0.58, so we use a multiplier of 1.7 to adjust all probabilities of death for those with AIDS who are not on treatment (1/0.58 = 1.7).[18] A set of calibration constants are also applied to all AIDS related deaths by age. For age buckets 15-29, 30-46, 50-64, and 65-100, we apply scalar multipliers of 2, 3, 1.75, and 1 respectively. This is done for calibration and can be explained as a reflection of local trends based on the AIDS death data.

#### 2e. Annual Probability of Infection

The following three properties drive new infections in the microsimulation: (1) PrEP and ART/Viral Suppression adherence by the individual and his partners, (2) Partnership patterns between subgroups, and (3) The number of HIV- and infectious HIV+ individuals in the population each given year. Table 5 outlines key parameters for determining annual probability of infection.

*Table 6: Infection properties*

| **Parameter** | **Value** | **Source** |
|---|---|---|
| Force of infection by race | [0.019, 0.0095, 0.0057] | Calibrated |
| Force of infection multiplier for ages 15-24 | 1.43 | Calibrated |
| PrEP adherence levels* (Proportion) | [0.2, 0.1, 0.7] | [19] |
| Relative risk of HIV infection by PrEP adherence level | [1, 0.42, 0.1] | [20–23] |

| | | |
|---|---|---|
| Treatment high adherence (Proportion) | 0.95 | [24] |
| Transmissibility multiplier if highly adherent treatment use | 0 | [24] |
| Number of partners for any individual by age group** | [8, 12, 12, 8, 12, 8, 0] | [25] |

** Levels or PrEP usage are low, medium, and high*

*** Age buckets are 15-19, 20-24, 25-34, 35-44, 45-54, 55-74, 75+*

*** Race groups are non-Hispanic Black, Hispanic, non-Hispanic White*

We consider three adherence levels for PrEP usage: 20% at low adherence, 10% at medium adherence, and 70% at high adherence[26]. At low adherence, PrEP is considered to have no effect, while at high adherence, 90% of PrEP users would be protected [20]. At medium adherence, 58% of the users would be protected[26]. Similarly, we consider individuals indicated to have viral suppression by the end of the year to have ART adherence levels that are either low (5% of treated individuals) or high (95% of treated individuals). Following information from the Partner2 study, high level users are considered not infectious while low level users remain infectious[24].

While there are multiple ways that HIV can be transmitted between MSM, the primary form of transmission is unprotected sexual contact. Models that are not stratified by age or race\ethnicity are unable to consider partnership patterns with respect to these characteristics and must assume purely non-assortative partnerships that assign equal likelihood for any individuals in the simulation to be partners, regardless of age or race/ethnicity. An alternative approach is to use empirical preferential partnership patterns (See section titled "Partnership Mixing Matrix"). In this framework, an individual's partnerships pairings depend on their own age- and race/ethnicity and that of their potential partner. In our simulation, we identify the likelihood of a transmissible contact and average number of sexual partners based on an individual's race/ethnicity (Black, Hispanic, White) and age (15-19 years, 20-24 years, 25-34 years, 35-44 years, 45-54 years, 55-74 years, 75-99 years) using data collected by a Los Angeles Lesbian, Gay, Bisexual, and Transgender (LGBT) Center. These probabilities, organized into a mixing/partnership matrix, are utilized to determine the annual probability of infection for different race/ethnicity and age groups each year, as the HIV positive population changes over time for each race/ethnicity and age group.

Finally, in determining the likelihood of new infection for an HIV negative individual, we only consider individuals who are not virally suppressed or have low adherence if virally suppressed to be infectious. The number of HIV positive individuals changes year to year as individuals move between health states -- new individuals can become virally suppressed, and previously virally suppressed individuals can fall out of viral suppression.

Based on these properties, the probability of infection for a susceptible individual is determined annually using the following characteristics: (1) individual's race/ethnicity and age, (2) race/ethnicity and age of partners, (3) average annual number of partnerships, (4) current population and the number of infectious individuals, (5) general ART adherence levels (6) individual's PrEP status and PrEP adherence levels. We do not explicitly differentiate between high- and low-risk MSM, as data on race/ethnicity on risky behavior is limited. We use different force of infection calibration constants to capture some of the differences seen between races. The full equation and variables for the annual probability of infection is discussed in the section titled "Equation Formulation".

#### 2e.i. Partnership Mixing Matrix

We model partnerships using a partnership matrix that captures the race/age preferences of individuals in the simulation. The matrix is informed from empirical data collected from an LA County LGBT Center Survey.[25] We identify three race/ethnicities (non-Hispanic Black, Hispanic, non-Hispanic White) and seven age ranges (15-19, 20-24, 25-34, 35-44, 45-54, 55-74, 75-99). Under these categorizations, we have 21 different demographic characterizations for which we define partnership preferences. Values in the matrix specify the probability an individual has partner of a specific race/ethnicity and age combination (column), conditioned on the race/ethnicity and age combination of that individual (row). By its construction, the mixing matrix is a square matrix where rows represent demographic characterization of the HIV negative individual, and the columns represent the demographic characterization of possible partners. Under this structure, the rows must sum to one, but the columns do not.

It is also important to consider the average annual number of partners an individual may have. Using survey responses regarding the number of partners in prior three months from the LA LGBT center data, we estimated that the number of partners an individual had by age where 8, 12, 12, 8, 12, and 8 for age groups 15-19, 20-24, 25-34, 35-44, 45-54, and 55-74 respectively. We assume no partnerships after age 75. We did not see differences occur by race/ethnicity with regards to number of partners.

#### 2.e.ii. Equation Formulation

The annual probability of infection for each demographic characterization is determined dynamically based on a set of time-invariant parameters, the current specified population size, and the current number of HIV positive individuals in the specified population. Calibration variables for the force of infection are also necessary in our dynamic formulation of annual probability of infection for a given demographic group.

Considering the demographic characterization of an individual and the associated partnership preference pattern, we determine the probability an individual might be get HIV by at least 1 of their partners each year. We present variable definitions and our probability of infection equation below.

*Equation 1: Probability of infection*

$$P(Infection) = (1 - \prod_{d_p \in D} (1 - \gamma \beta_d \frac{I_{dp}}{N_{dp}})^{P_d M_{dp}})\phi$$

$$\text{where} \qquad I_{dp} = I_{nt,dp} + I_{t,dp}(1-\alpha)$$

**Variable definitions**

$D$: set of possible demographic groups
$d$: demographic of the susceptible individual
$P_d$: number of partners an individual in that demographic group would have
$d_p$: demographic group of the partner
$M_{dp}$: probability of the susceptible individual mixing with the partner demographic group
$I_{dp}$: infected that can transmit in demographic group of partner
$I_{t,dp}$: infected group on treatment
$I_{nt,dp}$: infected group not on treatment
$N_{dp}$: population in demographic group of partner
$\phi$: PrEP adherence multiplier
$\alpha$: ART adherence proportion
$\beta_d$: Force of infection (different for each race/ethnicity)
$\gamma$: Force of infection multiplier for ages $\leq 24$

#### 3. Calibration

A hierarchical process was used to calibrate the microsimulation. First, calibration targets were identified from LA County surveillance data. We prioritized aggregate calibration targets over stratified targets (age, race or stage specific). We prioritize targets relating to new diagnosis over those pertaining to total diagnosed PLWH because our outcomes of interest are more related to new infections and new diagnosis than total PLWH. Further, among our stratified targets, race/ethnicity targets are prioritized over age related targets, which are prioritized over stage related targets. This is because of measures relating to race and age in the surveillance data are more likely to be accurate than stage data as HIV stage varies over time in a less predictable nature than a characteristic such as age. AIDS diagnosed deaths are our lowest priority among the stratified calibration targets.

In calibrating the model, we changed uncertain values (calibration parameters) to align model output with trends observed in the LA County Department of Public Health surveillance data. Uncertain input parameters that required calibration include attainment of viral suppression by race/ethnicity and age and disease progression while on and off treatment. Further, we also introduced three calibration constants: (1) a multiplier used to represent the force of infection in the annual probability of infection calculation (varies by race) because of differences by race in attributes that could impact risk of infection (such as STDs)[27,28], (2) a multiplier that scaled up the risk of infection among individuals under age 24, as this group has historically shown higher incidence rates and has been associated with risky behaviors including low testing rates, substance use, and low rates of condom use[29,30]. (3) a multiplier that adjusted AIDS related death probabilities, as these values were derived from national data and not specific to LA County. During the calibration process, we varied these inputs such that we attained model outputs that were consistent with observed data across several metrics simultaneously (calibration targets). All calibration targets used were annual counts determined from LA County surveillance data from 2012-2016[3].

In the hierarchical process, we first identify calibration parameter values that, as best as possible, satisfies meeting the associated calibration target within a fixed +/- 15% of the documented surveillance value. We use a large range in our assessment because of uncertainty in the surveillance data. The calibration parameter is held constant as calibration is done for the next target. If the new target is not attainable or requires modification of a previous calibration parameters, modifications are made to the prior calibration parameters such that prior calibration targets remain as close to satisfied as possible. In calibrating the model. Table 6 below depicts the calibration targets, parameters, and prioritization. Calibration parameters values have previously been presented in the prior subsections. Graphs are presented for each calibration target. We additionally report root mean squared values of the percentage error for each calibration target (over all calibration years). Values are typically below 15% for many targets. We find that our calibrated model outputs fall within 10% of root mean square error (RMSE) on percent error for the number of PLWH, new diagnoses, viral suppression, and deaths over the calibration period for the entire population. We accepted larger deviations for age, race, and HIV stage specific calibration targes as subgroup data often had small values (and a single case represented a larger percentage).

*Table 7: Calibration table*

| Priority Level | Calibration Targets | Calibration Parameter |
|---|---|---|
| High | New Diagnosis (Overall and AIDS Specific) | Calibration Constant for infections by race (Force of infection per interaction) |
| High | Diagnosed PLWH (Overall) | Calibration Constant for infections by race (Force of infection per interaction) |
| High | AIDS Diagnosed Deaths (Overall) | Calibration constant for all AIDS Deaths by different age buckets |
| High | Diagnosed PLWH on Treatment (Overall) | Attaining viral suppression by race and age |
| Medium | New Diagnosis by Race | Calibration Constant for infections by race (Force of infection per interaction) and attaining viral suppression by race and age |
| Medium | New Diagnosis by Age | Calibration multiplier for infections in younger population (<24 years old) |
| Medium | New Diagnosis by Stage | Transition between stages depending on viral suppression status |
| Medium | Diagnosed PLWH on Treatment by Race | Attaining viral suppression by race and age |
| Medium | Diagnosed PLWH on Treatment by Age | Attaining viral suppression by race and age |
| Medium | Diagnosed PLWH by Race | Attaining viral suppression by race and age and Calibration Constant for infections by race (Force of infection per interaction) |
| Medium | Diagnosed PLWH by Age | Calibration multiplier for infections in younger population (<24 years old) |
| Low | AIDS Diagnosed Deaths by Race | Calibration constant for all AIDS Deaths by different age buckets |
| Low | AIDS Diagnosed Deaths by Age | Calibration constant for all AIDS Deaths by different age buckets |

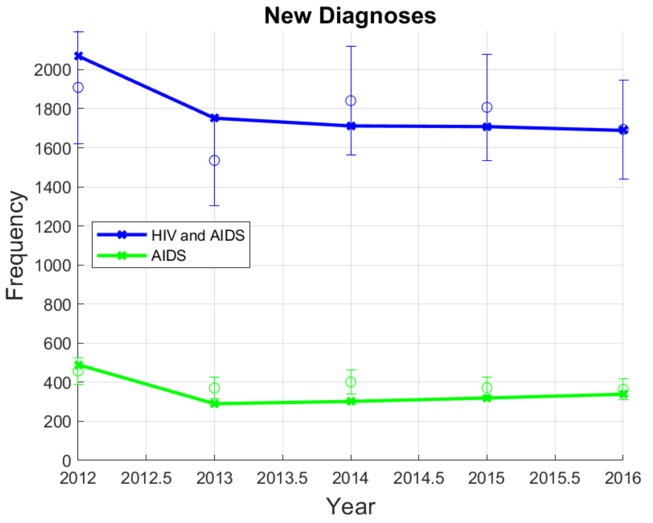

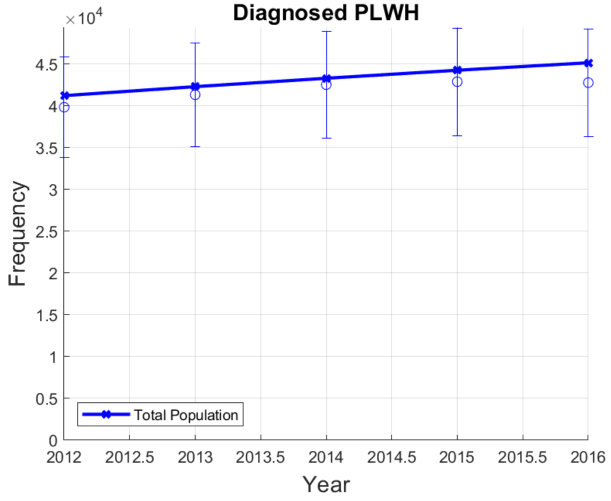

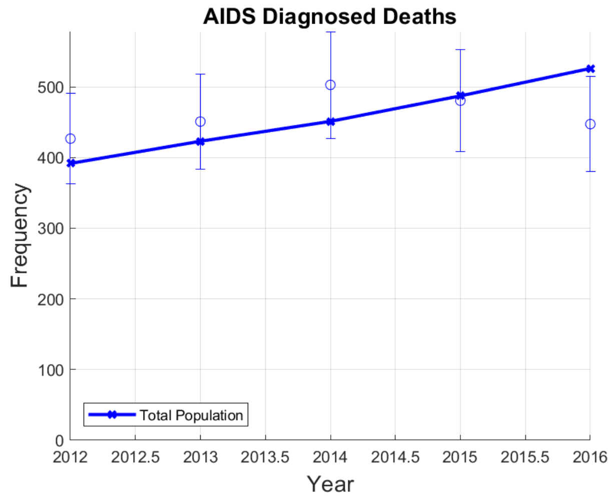
AIDS Diagnosed Deaths
Frequency
0
100
200
300
400
500
2012
2012.5
2013
2013.5
2014
2014.5
2015
2015.5
2016
Year
Total Population

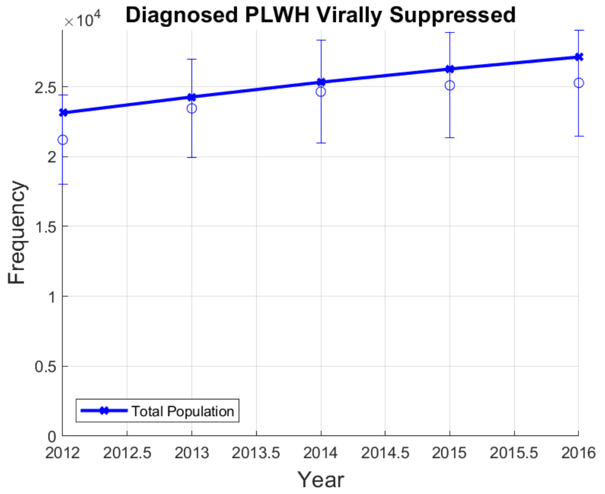
Diagnosed PLWH Virally Suppressed
×10^4
Frequency
0
0.5
1
1.5
2
2.5
2012
2012.5
2013
2013.5
2014
2014.5
2015
2015.5
2016
Year
Total Population

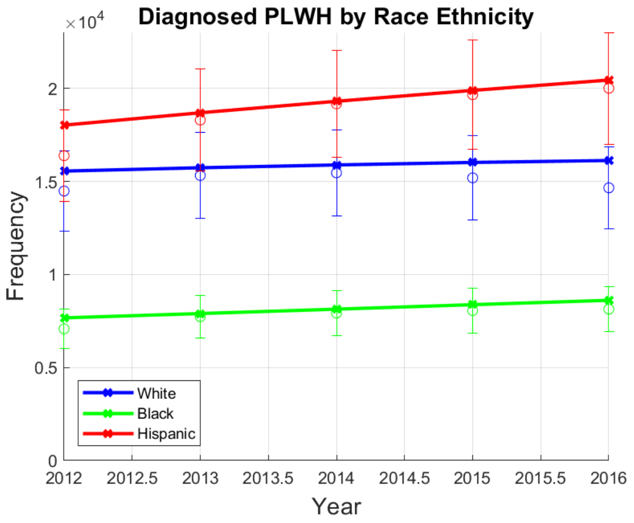
Diagnosed PLWH by Race Ethnicity
×10^4
Frequency
0
0.5
1
1.5
2
2012
2012.5
2013
2013.5
2014
2014.5
2015
2015.5
2016
Year
White
Black
Hispanic

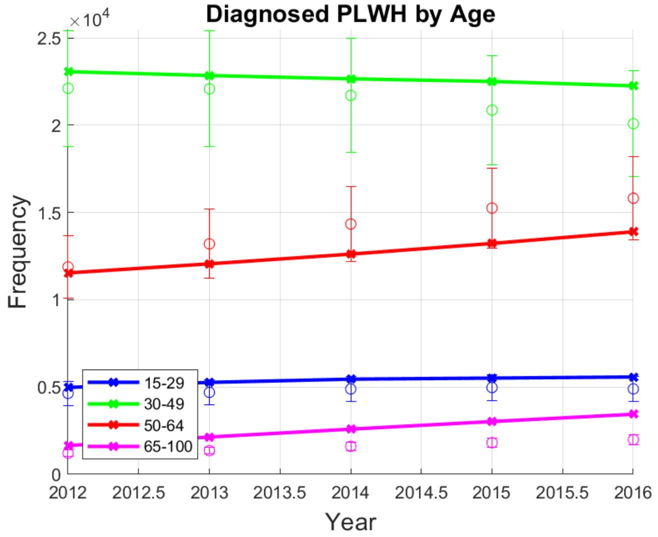
Diagnosed PLWH by Age
×10^4
Frequency
0
0.5
1
1.5
2
2.5
2012
2012.5
2013
2013.5
2014
2014.5
2015
2015.5
2016
Year
15-29
30-49
50-64
65-100

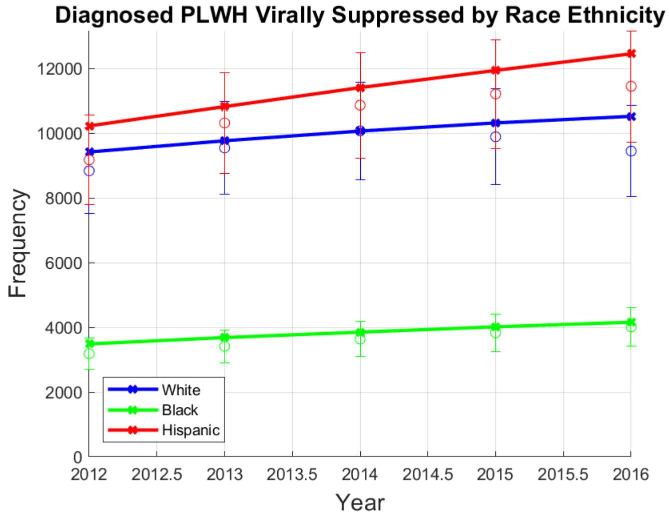
Diagnosed PLWH Virally Suppressed by Race Ethnicity
Frequency
0
2000
4000
6000
8000
10000
12000
2012
2012.5
2013
2013.5
2014
2014.5
2015
2015.5
2016
Year
White
Black
Hispanic

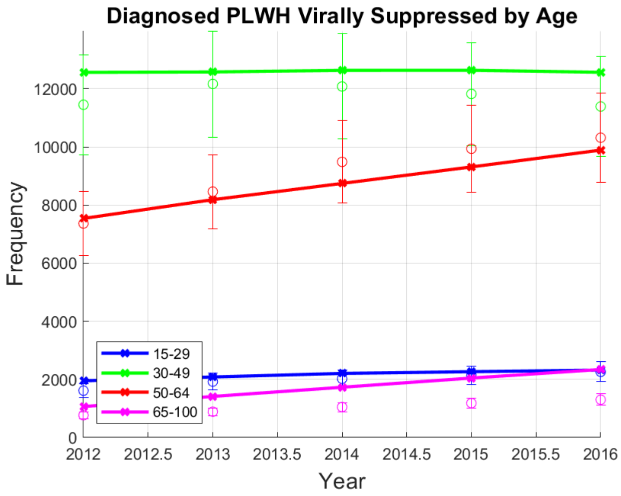
Diagnosed PLWH Virally Suppressed by Age
Frequency
0
2000
4000
6000
8000
10000
12000
2012
2012.5
2013
2013.5
2014
2014.5
2015
2015.5
2016
Year
15-29
30-49
50-64
65-100

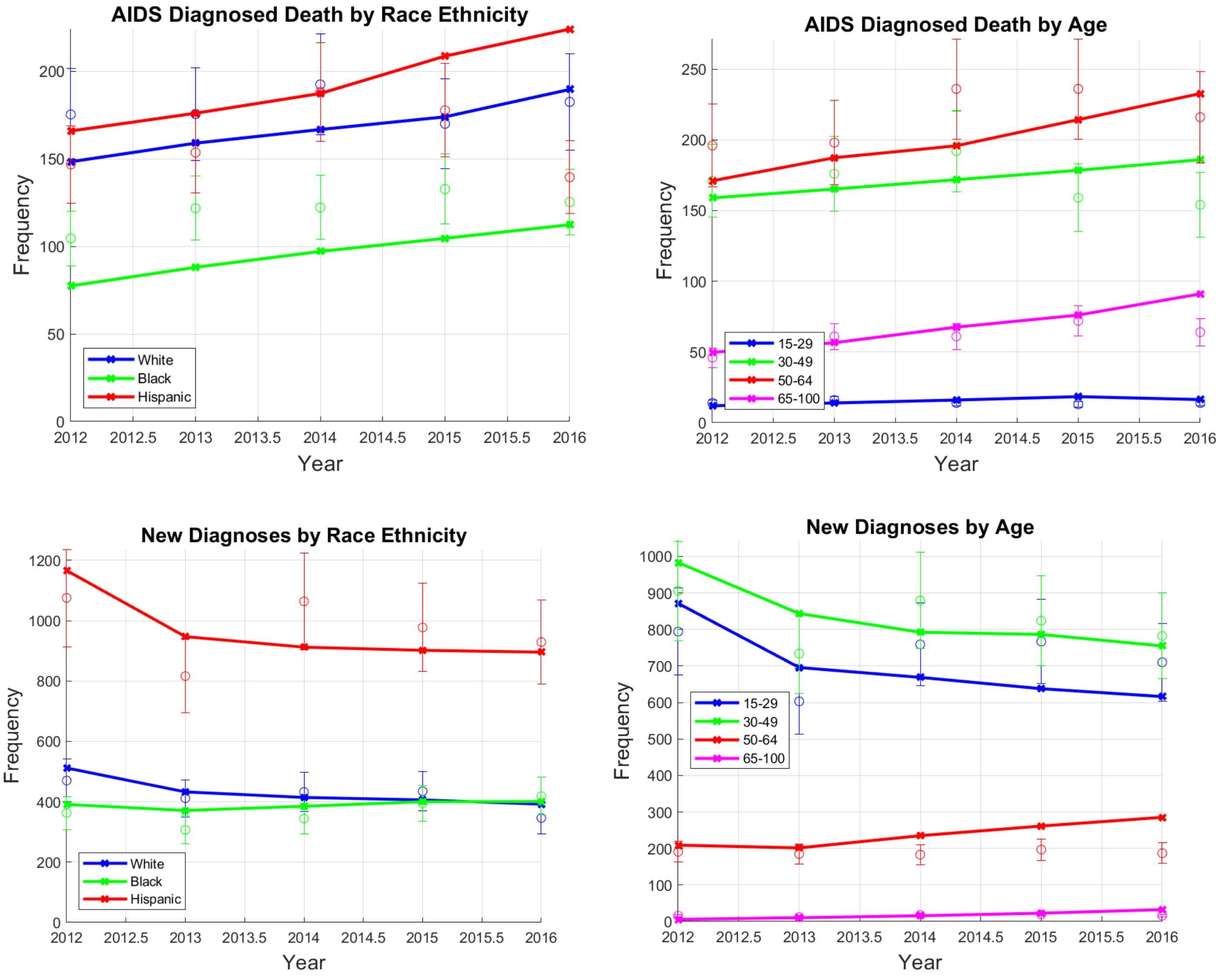

*Figure 1: All calibration graphs*

*Table 8: RMSE for Calibration parameters. RMSE is calculated for aggregate target as well as stratified targets*

**RMSE for Calibration Targets**

| Stratification | PLWH | New Diagnoses | VLS | Deaths |
|---|---|---|---|---|
| **Overall** | 0.035 | 0.090 | 0.060 | 0.084 |
| ***Age*** | | | | |
| 15-29 | 0.114 | 0.132 | 0.112 | 0.229 |
| 30-49 | 0.068 | 0.088 | 0.075 | 0.125 |
| 50-64 | 0.105 | 0.292 | 0.052 | 0.112 |
| 65-100 | 0.602 | 0.536 | 0.647 | 0.203 |
| ***Race/Ethnicity*** | | | | |
| White | 0.063 | 0.084 | 0.063 | 0.102 |
| Black | 0.051 | 0.117 | 0.067 | 0.220 |
| Hispanic | 0.047 | 0.100 | 0.077 | 0.295 |
| ***HIV Stage*** | | | | |
| CD4 >= 500 | NA | 0.101 | NA | NA |
| 200 <= CD4 <= 499 | NA | 0.132 | NA | NA |
| CD4 <= 200 | NA | 0.168 | NA | NA |

#### 4. Validation

The *Los Angeles County HIV/AIDS Strategy for 2020 and Beyond* report was used for internal validation of the model for counts on undiagnosed PLWH, viral suppression, new diagnoses, and total PLWH in 2016[13]. The *HIV Surveillance Annual Report 2019* is used to internally validate race/ethnicity related difference in LA County in terms of diagnosis rates, incidence rates, HIV status awareness, and viral suppression.[31] To externally validate prevalence and incidence outcomes, we used data from the CDC fact sheet for HIV Among Gay and Bisexual Men adjusted for differences between LAC and the national level in the proportion of new diagnoses among MSM.[32] While this can add additional uncertainty, it allows us to benchmark our model outcomes to nationally reported outcomes. We also validated PrEP coverage outcomes on estimates from Sullivan that examine national PrEP trends in the United States[33] and estimates from AIDSVu for PrEP usage in LAC.[34]

Internal validation values identified from the reports are for the entire LA County population, not MSM specifically. To account for this, we scale the values by 0.84 (if the value is a count) based on an estimated 84% of the HIV positive population in LA being reported as MSM per the report[13].

Internal validation measures at the stratified by race/ethnicity for diagnosis rates, incidence rates, HIV status awareness and viral suppression are either proportions or rates. If the validation value is a proportion, we assume that the MSM proportion is the same as the county proportion. If the validation measure is a rate, we use a relative rate (relative to Hispanic), to determine a value for comparison.

External validation for incidence and prevalence rates (per 100,000) use an estimated national MSM population count of 4,503,800 as reported by Grey[2]. However, at the national level, MSM only account for ~62% of the HIV positive population while this count is ~84% in LA[13,32]. We therefore scale the calculated rates by 1.35 (84/62) to account for difference in expected incidence for LA county.

For all validation, we apply +/- 10% deviations to determine ranges when only single values were presented. We present the computations to determine the values and ranges used in our validation table in the main manuscript. We then present the simulated values for each validation target.

*Table 9: Validation Ranges*

| **INTERNAL VALIDATION** | |
|---|---|
| **Undiagnosed PLWH (2016)** [13] | |
| *Min Calculation* | $8654 * 0.84 * 0.90 = 6542.424 \approx 6500$ |
| *Max Calculation* | $8654 * 0.84 * 1.10 = 7996.296 \approx 8000$ |
| **Percent Undiagnosed PLWH (2016)** [13] | |
| *Min Calculation* | $\frac{8654}{60946} * 0.90 * 100 = 12.8 \approx 13$ |
| *Max Calculation* | $\frac{8654}{60946} * 1.10 * 100 = 15.6 \approx 16$ |
| **Percent Virally Suppressed (2016)** [13] | |
| *Min Calculation* | $60 * 0.90 = 54$ |
| *Max Calculation* | $60 * 1.10 = 66$ |
| **New Diagnosis (2016)** [13] | |
| *Min Calculation* | $1700 * 0.84 = 1470$ |
| *Max Calculation* | $2000 * 0.84 = 1680$ |
| **Estimated PLWH (2016)** [13] | |

| | |
|---|---|
| *Min Calculation* | $59446 * 0.84 * 0.90 = 44941.18 \approx 45000$ |
| *Max Calculation* | $59446 * 0.84 * 1.10 = 54928.1 \approx 55000$ |
| **Diagnosis Rate Black Relative to Hispanic (2018)** [31] | |
| *Min Calculation* | $\frac{91}{37} * 0.90 * 100 = 2.21$ |
| *Max Calculation* | $\frac{91}{37} * 1.10 * 100 = 2.70$ |
| **Diagnosis Rate White Relative to Hispanic (2018)** [31] | |
| *Min Calculation* | $\frac{23}{37} * 0.90 * 100 = 0.56$ |
| *Max Calculation* | $\frac{23}{37} * 1.10 * 100 = 0.68$ |
| **Incidence Rate Black Relative to Hispanic (2018)** [31] | |
| *Min Calculation* | $\frac{54}{21} * 0.90 * 100 = 2.31$ |
| *Max Calculation* | $\frac{54}{21} * 1.10 * 100 = 2.83$ |
| **Incidence Rate White Relative to Hispanic (2018)** [31] | |
| *Min Calculation* | $\frac{12}{21} * 0.90 * 100 = 0.51$ |
| *Max Calculation* | $\frac{12}{21} * 1.10 * 100 = 0.63$ |
| **Proportion of Black MSM PLWH Aware of HIV Status (2018)** [31] | |
| *Min Calculation* | $0.74 * 0.90 = 0.66$ |
| *Max Calculation* | $0.74 * 1.10 = 0.81$ |
| **Proportion of Hispanic MSM PLWH Aware of HIV Status (2018)** [31] | |
| *Min Calculation* | $0.77 * 0.90 = 0.69$ |
| *Max Calculation* | $0.77 * 1.10 = 0.85$ |
| **Proportion of White MSM PLWH Aware of HIV Status (2018)** [31] | |
| *Min Calculation* | $1 * 0.90 = 0.90$ |
| *Max Calculation* | $1 * 1.10 = 1.10$ |
| **Proportion of Black PLWH Virally Suppressed (2018)** [31] | |
| *Min Calculation* | $0.55 * 0.90 = 0.50$ |
| *Max Calculation* | $0.55 * 1.10 = 0.61$ |
| **Proportion of Hispanic PLWH Virally Suppressed (2018)** [31] | |
| *Min Calculation* | $0.61 * 0.90 = 0.56$ |
| *Max Calculation* | $0.61 * 1.10 = 0.67$ |
| **Proportion of White PLWH Virally Suppressed (2018)** [31] | |
| *Min Calculation* | $0.62 * 0.90 = 0.56$ |
| *Max Calculation* | $0.62 * 1.10 = 0.68$ |
| **EXTERNAL VALIDATION** | |
| **Incidence Rate per 100,000 (2016)** [2,13,32] | |
| *Min Calculation* | $\frac{26400 + 1200}{4503800 - 648500 - 58600} * 100000 * \frac{84}{62} * 0.90 = 886.4 \approx 890$ |
| *Max Calculation* | $\frac{26400 + 1200}{4503800 - 648500 - 58600} * 100000 * \frac{84}{62} * 1.10 = 1083.4 \approx 1100$ |
| **PLWH Rate pre 100,000 (2016)** [2,13,32] | |
| *Min Calculation* | $\frac{648500 + 58600}{4503800} * 100000 * \frac{84}{62} * 0.90 = 19144 \approx 19100$ |

| | |
|---|---|
| *Max Calculation* | $\frac{648500 + 58600}{4503800} * 100000 * \frac{84}{62} * 1.10 = 23398 \approx 23400$ |
| **Percent PrEP Coverage in 2017** [33] | |
| No calculation was needed for the PrEP coverage range. The range cited in literature is for total proportion of population that has used PrEP ever. Because we only determine PrEP coverage for a given year, we expect our value to be on the lower end of this metric because discontinuation rates for PrEP are high. | |

*Table 10: Model Validation: If literature only contains a single value, lower bound (LB) and upper bound (UB) are 10% deviations. If literature only contains an upper and lower bound, the value is the value is the mean.*

| Outcome | Literature Value [LB, UB] | Simulated Value (Std) | Source |
|---|---|---|---|
| Undiagnosed PLWH (2016) | 7269 [6500, 8000] | 6960 [6930, 6990] | [13] |
| Percent Undiagnosed PLWH (2016) | 14 [13, 16] | 13.3 [13.2, 13.4] | [13] |
| Percent Virally Suppressed (2016) | 60 [54, 66] | 60 [59.9-60.1] | [13] |
| New Diagnosis (2016) | 1575 [1470, 1680] | 1689 [1672, 1706] | [13] |
| Estimated PLWH (2016) | 49935 [45000, 55000] | 52136 [52090, 52182] | [13] |
| Estimated PrEP users (2018) | 8350 | 6878 [6849, 6907] | [35] |
| Diagnosis Rate Black relative to Hispanic (2018) | 2.45 [2.205, 2.695] | 2.83 [2.76, 2.89] | [31] |
| Diagnosis Rate White relative to Hispanic (2018) | 0.62 [0.56, 0.68] | 0.67 [0.66, 0.68] | [31] |
| Incidence Rate Black relative to Hispanic (2018) | 2.57 [2.31, 2.83] | 2.59 [2.54, 2.64] | [31] |
| Incidence Rate White relative to Hispanic (2018) | 0.57 [0.51, 0.63] | 0.61 [0.60, 0.62] | [31] |
| Black MSM Aware of HIV Status (2018) | 0.74 [0.66, 0.81] | 0.846 [0.845, 0.847] | [31] |
| Hispanic MSM Aware of HIV Status (2018) | 0.77 [0.69, 0.85] | 0.858 [0.857, 0.859] | [31] |
| White MSM Aware of HIV Status (2018) | 1 | 0.889 [0.888, 0.889] | [31] |
| Percent Black PLWH Virally Suppressed | 0.55 [0.50, 0.61] | 0.492 [0.491, 0.494] | [31] |
| Percent Hispanic PLWH Virally Suppressed | 0.61 [0.56, 0.67] | 0.622 [0.621, 0.624] | [31] |
| Percent White PLWH Virally Suppressed | 0.62 [0.56, 0.68] | 0.666 [0.665, 0.667] | [31] |
| Overall Incidence Rate per 100,000 (2016) | 985 [890, 1100] | 968 [960, 977] | [2,13,32] |
| PLWH Rate per 100,000 (2016) | 21271 [19100, 23400] | 20291 [20274, 20308] | [2,13,32] |
| Percent PrEP Coverage (2017) | 5.5 [2, 9] | 2.62 [2.61, 2.63] | [33] |

#### 5. Policies

The policies implemented allocated different quantities of PrEP to different race groups. For the single race/ethnicity policies, all PrEP is allocated to the single race/ethnicity group. For the distributed policies, the PrEP is spread among the race/ethnicity groups. In the table below, we present the specific quantities of PrEP allocated to each race/ethnicity group under different allocation schemes. We test our polices at the 3000, 6000, and 9000 additional annual prescriptions levels. We restrict the maximum annual PrEP increase to 9000 because higher values will result in an oversaturation of PrEP for the Black MSM group under certain allocation schemes by the end of the simulated intervention period, making these scenarios unsuitable for comparison with others.

The Equal allocation scheme distributes PrEP equally to each racial/ethnic group. The count policy distributes proportionally to racial distribution of HIV among PLWH (21% non-Hispanic Black, 47% Hispanic, 32% non-Hispanic White). The rate policy distributes proportionally to the new diagnosis rates in each race/ethnicity group (63% non-Hispanic Black, 24% Hispanic, 13% non-Hispanic White). These approximations align with the total PLWH by race in 2018 and new diagnoses by race in 2016 outlined in the 2018 LAC HIV surveillance report.[36]

*Table 11: Scenario proportion details are as follows: Black (100% to non-Hispanic Black), Hispanic (100% to Hispanics), White (100% to non-Hispanic Whites), Equal (33% to each of the three race/ethnicities), Count (21% non-Hispanic Black, 47% Hispanic, and 32% non-Hispanic White), Rate (63% non-Hispanic Black, 24% Hispanic, 13% non-Hispanic White). Note that an additional 1000 PrEP is approximately a 20%-22% increase in PrEP uptake relative to the baseline uptake in 2020. All allocation schemes are tested at the 3000, 6000, and 9000 additional annual PrEP coverage levels.*

| | Reference Name | Allocation Scheme |
|---|---|---|
| **Single Race Policies** | Black | Only non-Hispanic Black |
| | Hispanic | Only Hispanic |
| | White | Only non-Hispanic White |
| **Distributed Policies** | Equal | Equal quantity of PrEP for each race/ethnicity |
| | Count | Proportional by race/ethnicity based on number of PLWH in each race/ethnicity |
| | Rate | Proportional by race/ethnicity based on new diagnosis rate in each race/ethnicity |

*Table 12: PrEP distribution breakdown for all distributed policies*

| Total PrEP Count | Allocation Name | Black | Hispanic | White |
|---|---|---|---|---|
| 3000 | Equal | 1000 | 1000 | 1000 |
| 6000 | Equal | 2000 | 2000 | 2000 |
| 9000 | Equal | 3000 | 3000 | 3000 |
| 3000 | Count | 632 | 1421 | 947 |
| 6000 | Count | 1263 | 2842 | 1895 |

| | | | | |
|---|---|---|---|---|
| 9000 | Count | 1895 | 4263 | 2842 |
| 3000 | Rate | 1883 | 726 | 391 |
| 6000 | Rate | 3766 | 1452 | 781 |
| 9000 | Rate | 5649 | 2179 | 1172 |

#### 6. Standard Errors on Cumulative Infections Averted

In Figure 3 of the main manuscript, we presented cumulative infections averted for the 3000, 6000, and 9000 PrEP coverage levels for all allocation schemes. Presented in table 12 are the standard errors associated with the total cumulative infections averted.

*Table 13: Standard Errors on Cumulative Infections Averted*

| | 3000 | | | | 6000 | | | | 9000 | | | |
|---|---|---|---|---|---|---|---|---|---|---|---|---|
| | *Mean* | *StdError* | *95% CI (LB)* | *95% CI (UB)* | *Mean* | *StdError* | *95% CI (LB)* | *95% CI (UB)* | *Mean* | *StdError* | *95% CI (LB)* | *95% CI (UB)* |
| **Black** | 1019.5 | 85.4 | 852.2 | 1186.8 | 2185.6 | 81.2 | 2026.5 | 2344.7 | 3143.2 | 82.2 | 2982.1 | 3304.3 |
| **Hispanic** | 320.3 | 81.1 | 161.4 | 479.3 | 896.0 | 80.1 | 739.0 | 1053.0 | 1420.0 | 76.4 | 1270.2 | 1569.7 |
| **White** | 336.5 | 82.5 | 174.7 | 498.2 | 580.1 | 84.5 | 414.5 | 745.8 | 938.9 | 88.2 | 766.0 | 1111.9 |
| **Equal** | 669.4 | 78.3 | 515.9 | 822.9 | 1265.8 | 74.2 | 1120.3 | 1411.3 | 1869.4 | 84.1 | 1704.5 | 2034.4 |
| **Count** | 534.9 | 93.0 | 352.6 | 717.2 | 1159.2 | 84.6 | 993.5 | 1325.0 | 1695.9 | 75.1 | 1548.6 | 1843.2 |
| **Rate** | 753.8 | 91.2 | 574.9 | 932.6 | 1648.5 | 85.5 | 1480.9 | 1816.2 | 2503.8 | 85.6 | 2335.9 | 2671.6 |

#### 7. Measuring Equality in 2035

To assess disparities after 15 years of the policies being implemented (2021-2035), we calculate the Gini index using incidence rate in 2035. Other health studies on disparities use QALYs generated or other measures of overall health, but we focus on new cases as a more proximal measure of HIV burden. Other measures commonly used are Atkinson index and Kolm index. We present these values alongside the Gini index for all allocation strategies and all PrEP levels tested.

*Table 14: Equality Measures for polices at the 9000 PrEP level*

| | No Policy | Black | Hispanic | White | Equal | Count | Rate |
|---|---|---|---|---|---|---|---|
| **Gini Index** | 0.24 | 0.13 | 0.24 | 0.27 | 0.21 | 0.22 | 0.17 |
| **Atkinson ($\varepsilon = 1$)** | 0.11 | 0.04 | 0.11 | 0.14 | 0.08 | 0.1 | 0.06 |
| **Atkinson ($\varepsilon = 7$)** | 0.34 | 0.24 | 0.3 | 0.42 | 0.32 | 0.33 | 0.27 |
| **Atkinson ($\varepsilon = 30$)** | 0.42 | 0.34 | 0.39 | 0.5 | 0.4 | 0.41 | 0.36 |
| **Kolm ($\alpha = 0.025$)** | 252.70 | 156.86 | 205.16 | 279.61 | 210.09 | 214.84 | 176.87 |
| **Kolm ($\alpha = 0.5$)** | 292.16 | 196.77 | 244.61 | 318.92 | 249.69 | 254.4 | 216.62 |

*Table 15: Equality Measures for polices at the 6000 PrEP level*

| | No Policy | Black | Hispanic | White | Equal | Count | Rate |
|---|---|---|---|---|---|---|---|
| **Gini Index** | 0.24 | 0.16 | 0.23 | 0.26 | 0.22 | 0.23 | 0.2 |
| **Atkinson ($\varepsilon = 1$)** | 0.11 | 0.05 | 0.11 | 0.12 | 0.09 | 0.1 | 0.07 |
| **Atkinson ($\varepsilon = 7$)** | 0.34 | 0.27 | 0.31 | 0.39 | 0.32 | 0.33 | 0.3 |
| **Atkinson ($\varepsilon = 30$)** | 0.42 | 0.36 | 0.39 | 0.47 | 0.4 | 0.41 | 0.38 |
| **Kolm ($\alpha = 0.025$)** | 252.70 | 181.27 | 217.65 | 269.94 | 218.5 | 227.47 | 200.88 |
| **Kolm ($\alpha = 0.5$)** | 292.16 | 221.06 | 257.17 | 309.29 | 258.06 | 267.03 | 240.55 |

*Table 16: Equality Measures for polices at the 3000 PrEP level*

| | No Policy | Black | Hispanic | White | Equal | Count | Rate |
|---|---|---|---|---|---|---|---|
| **Gini Index** | 0.24 | 0.20 | 0.24 | 0.25 | 0.23 | 0.23 | 0.22 |
| **Atkinson ($\varepsilon = 1$)** | 0.11 | 0.08 | 0.11 | 0.12 | 0.1 | 0.1 | 0.09 |
| **Atkinson ($\varepsilon = 7$)** | 0.34 | 0.31 | 0.32 | 0.38 | 0.34 | 0.33 | 0.32 |
| **Atkinson ($\varepsilon = 30$)** | 0.42 | 0.40 | 0.40 | 0.46 | 0.42 | 0.41 | 0.4 |
| **Kolm ($\alpha = 0.025$)** | 252.70 | 217.20 | 238.16 | 268.45 | 239.86 | 236.67 | 226.77 |
| **Kolm ($\alpha = 0.5$)** | 292.16 | 256.80 | 277.65 | 307.85 | 279.36 | 276.14 | 266.33 |

Gini Index, Atkinson Index, and Kolm Index are calculated using the following definitions:

**Gini Index[37,38]:**

$$G = \frac{\sum_i^n \sum_j^n |x_i - x_j|}{2n^2\bar{x}}$$

$$G = \frac{2}{n^2\bar{x}} \sum_i^n i(x_i - \bar{x})$$

$$G = \frac{\sum_i^n (2i - n - 1)x_i}{n\sum_i^n x_i}$$

where $x_i$ is the incidence rate, n is the number of susceptible (HIV-) individuals, and *i* is the rank of values in ascending order:

**Atkinson Index[39]:**

$$A(\varepsilon) = 1 - \frac{[\sum_i^n f_i(y_i^{1-\varepsilon})]^{\frac{1}{1-\varepsilon}}}{\bar{y}} \; for \; 0 \leq \varepsilon \neq 1$$

$$A(\varepsilon) = 1 - \frac{\prod_i^N (y_i^{f_i})}{\bar{y}} \; for \; \varepsilon = 1$$

$f_i = \frac{w_i}{N}$, where $w_i$ is the number of people in subgroup $i$ and $N$ is the number of susceptible populations. $y_i$ is the incidence rate of race group $i$ and $\varepsilon$ is the parameter of inequality aversion

**Kolm Index:**

$$K(\alpha) = \frac{1}{\alpha}\log\left(\sum_{i}^{n} e^{\alpha(\bar{x}-x_i)} f_i\right)$$

$f_i = \frac{w_i}{N}$, where $w_i$ is the number of people in subgroup $i$ and $N$ is the number of susceptible populations. $y_i$ is the incidence rate of race group $i$ and $\alpha$ is the nonnegative parameter of inequality aversion

#### 8. Sensitivity Analysis

We perform sensitivity analysis on our partnership mixing matrix. In the current model, the partnership matrix is derived from empirically collected data at an LA LGBT Center. We recognize that this matrix may tend to have some biases associated with the population that visited the clinic at which the survey was administered. To see if the partnership matrix plays a significant impact on outcomes, we tested two other partnership matrix structure: assortative and uniform.

We define an assortative partnership matrix as a matrix where each race, only has partner preferences within their own race with equal probability for all age groups. A uniform partnership matrix is defined as when an individual has equal preferences for all other age/race groups. In all partnership matrices, we assume that ages 75+ no longer have partners and cannot become HIV positive. Presented in the figures below are heatmaps of the assortative and uniform partnership matrices.

| | | | Partner | | | | | | | | | | | | | | | | | | | | |
|---|---|---|---|---|---|---|---|---|---|---|---|---|---|---|---|---|---|---|---|---|---|---|---|
| | | Race | Non-Hispanic Black | | | | | | | Hispanic | | | | | | | Non-Hispanic White | | | | | | |
| | Race | Age | 15-19 | 20-24 | 25-34 | 35-44 | 45-54 | 55-74 | 75-99 | 15-19 | 20-24 | 25-34 | 35-44 | 45-54 | 55-74 | 75-99 | 15-19 | 20-24 | 25-34 | 35-44 | 45-54 | 55-74 | 75-99 |
| Susceptible Individual | Non-Hispanic Black | 15-19 | 0.17 | 0.17 | 0.17 | 0.17 | 0.17 | 0.17 | 0 | 0 | 0 | 0 | 0 | 0 | 0 | 0 | 0 | 0 | 0 | 0 | 0 | 0 | 0 |
| | | 20-24 | 0.17 | 0.17 | 0.17 | 0.17 | 0.17 | 0.17 | 0 | 0 | 0 | 0 | 0 | 0 | 0 | 0 | 0 | 0 | 0 | 0 | 0 | 0 | 0 |
| | | 25-34 | 0.17 | 0.17 | 0.17 | 0.17 | 0.17 | 0.17 | 0 | 0 | 0 | 0 | 0 | 0 | 0 | 0 | 0 | 0 | 0 | 0 | 0 | 0 | 0 |
| | | 35-44 | 0.17 | 0.17 | 0.17 | 0.17 | 0.17 | 0.17 | 0 | 0 | 0 | 0 | 0 | 0 | 0 | 0 | 0 | 0 | 0 | 0 | 0 | 0 | 0 |
| | | 45-54 | 0.17 | 0.17 | 0.17 | 0.17 | 0.17 | 0.17 | 0 | 0 | 0 | 0 | 0 | 0 | 0 | 0 | 0 | 0 | 0 | 0 | 0 | 0 | 0 |
| | | 55-74 | 0.17 | 0.17 | 0.17 | 0.17 | 0.17 | 0.17 | 0 | 0 | 0 | 0 | 0 | 0 | 0 | 0 | 0 | 0 | 0 | 0 | 0 | 0 | 0 |
| | | 75-79 | 0 | 0 | 0 | 0 | 0 | 0 | 0 | 0 | 0 | 0 | 0 | 0 | 0 | 0 | 0 | 0 | 0 | 0 | 0 | 0 | 0 |
| | Hispanic | 15-19 | 0 | 0 | 0 | 0 | 0 | 0 | 0 | 0.17 | 0.17 | 0.17 | 0.17 | 0.17 | 0.17 | 0 | 0 | 0 | 0 | 0 | 0 | 0 | 0 |
| | | 20-24 | 0 | 0 | 0 | 0 | 0 | 0 | 0 | 0.17 | 0.17 | 0.17 | 0.17 | 0.17 | 0.17 | 0 | 0 | 0 | 0 | 0 | 0 | 0 | 0 |
| | | 25-34 | 0 | 0 | 0 | 0 | 0 | 0 | 0 | 0.17 | 0.17 | 0.17 | 0.17 | 0.17 | 0.17 | 0 | 0 | 0 | 0 | 0 | 0 | 0 | 0 |
| | | 35-44 | 0 | 0 | 0 | 0 | 0 | 0 | 0 | 0.17 | 0.17 | 0.17 | 0.17 | 0.17 | 0.17 | 0 | 0 | 0 | 0 | 0 | 0 | 0 | 0 |
| | | 45-54 | 0 | 0 | 0 | 0 | 0 | 0 | 0 | 0.17 | 0.17 | 0.17 | 0.17 | 0.17 | 0.17 | 0 | 0 | 0 | 0 | 0 | 0 | 0 | 0 |
| | | 55-74 | 0 | 0 | 0 | 0 | 0 | 0 | 0 | 0.17 | 0.17 | 0.17 | 0.17 | 0.17 | 0.17 | 0 | 0 | 0 | 0 | 0 | 0 | 0 | 0 |
| | | 75-79 | 0 | 0 | 0 | 0 | 0 | 0 | 0 | 0 | 0 | 0 | 0 | 0 | 0 | 0 | 0 | 0 | 0 | 0 | 0 | 0 | 0 |
| | Non-Hispanic White | 15-19 | 0 | 0 | 0 | 0 | 0 | 0 | 0 | 0 | 0 | 0 | 0 | 0 | 0 | 0 | 0.17 | 0.17 | 0.17 | 0.17 | 0.17 | 0.17 | 0 |
| | | 20-24 | 0 | 0 | 0 | 0 | 0 | 0 | 0 | 0 | 0 | 0 | 0 | 0 | 0 | 0 | 0.17 | 0.17 | 0.17 | 0.17 | 0.17 | 0.17 | 0 |
| | | 25-34 | 0 | 0 | 0 | 0 | 0 | 0 | 0 | 0 | 0 | 0 | 0 | 0 | 0 | 0 | 0.17 | 0.17 | 0.17 | 0.17 | 0.17 | 0.17 | 0 |
| | | 35-44 | 0 | 0 | 0 | 0 | 0 | 0 | 0 | 0 | 0 | 0 | 0 | 0 | 0 | 0 | 0.17 | 0.17 | 0.17 | 0.17 | 0.17 | 0.17 | 0 |
| | | 45-54 | 0 | 0 | 0 | 0 | 0 | 0 | 0 | 0 | 0 | 0 | 0 | 0 | 0 | 0 | 0.17 | 0.17 | 0.17 | 0.17 | 0.17 | 0.17 | 0 |
| | | 55-74 | 0 | 0 | 0 | 0 | 0 | 0 | 0 | 0 | 0 | 0 | 0 | 0 | 0 | 0 | 0.17 | 0.17 | 0.17 | 0.17 | 0.17 | 0.17 | 0 |
| | | 75-99 | 0 | 0 | 0 | 0 | 0 | 0 | 0 | 0 | 0 | 0 | 0 | 0 | 0 | 0 | 0 | 0 | 0 | 0 | 0 | 0 | 0 |

| | | | Partner | | | | | | | | | | | | | | | | | | | | |
|---|---|---|---|---|---|---|---|---|---|---|---|---|---|---|---|---|---|---|---|---|---|---|---|
| | | Race | Non-Hispanic Black | | | | | | | Hispanic | | | | | | | Non-Hispanic White | | | | | | |
| | Race | Age | 15-19 | 20-24 | 25-34 | 35-44 | 45-54 | 55-74 | 75-99 | 15-19 | 20-24 | 25-34 | 35-44 | 45-54 | 55-74 | 75-99 | 15-19 | 20-24 | 25-34 | 35-44 | 45-54 | 55-74 | 75-99 |
| Susceptible Individual | Non-Hispanic Black | 15-19 | 0.06 | 0.06 | 0.06 | 0.06 | 0.06 | 0.06 | 0.00 | 0.06 | 0.06 | 0.06 | 0.06 | 0.06 | 0.06 | 0.00 | 0.06 | 0.06 | 0.06 | 0.06 | 0.06 | 0.06 | 0.00 |
| | | 20-24 | 0.06 | 0.06 | 0.06 | 0.06 | 0.06 | 0.06 | 0.00 | 0.06 | 0.06 | 0.06 | 0.06 | 0.06 | 0.06 | 0.00 | 0.06 | 0.06 | 0.06 | 0.06 | 0.06 | 0.06 | 0.00 |
| | | 25-34 | 0.06 | 0.06 | 0.06 | 0.06 | 0.06 | 0.06 | 0.00 | 0.06 | 0.06 | 0.06 | 0.06 | 0.06 | 0.06 | 0.00 | 0.06 | 0.06 | 0.06 | 0.06 | 0.06 | 0.06 | 0.00 |
| | | 35-44 | 0.06 | 0.06 | 0.06 | 0.06 | 0.06 | 0.06 | 0.00 | 0.06 | 0.06 | 0.06 | 0.06 | 0.06 | 0.06 | 0.00 | 0.06 | 0.06 | 0.06 | 0.06 | 0.06 | 0.06 | 0.00 |
| | | 45-54 | 0.06 | 0.06 | 0.06 | 0.06 | 0.06 | 0.06 | 0.00 | 0.06 | 0.06 | 0.06 | 0.06 | 0.06 | 0.06 | 0.00 | 0.06 | 0.06 | 0.06 | 0.06 | 0.06 | 0.06 | 0.00 |
| | | 55-74 | 0.06 | 0.06 | 0.06 | 0.06 | 0.06 | 0.06 | 0.00 | 0.06 | 0.06 | 0.06 | 0.06 | 0.06 | 0.06 | 0.00 | 0.06 | 0.06 | 0.06 | 0.06 | 0.06 | 0.06 | 0.00 |
| | | 75-79 | 0.00 | 0.00 | 0.00 | 0.00 | 0.00 | 0.00 | 0.00 | 0.00 | 0.00 | 0.00 | 0.00 | 0.00 | 0.00 | 0.00 | 0.00 | 0.00 | 0.00 | 0.00 | 0.00 | 0.00 | 0.00 |
| | Hispanic | 15-19 | 0.06 | 0.06 | 0.06 | 0.06 | 0.06 | 0.06 | 0.00 | 0.06 | 0.06 | 0.06 | 0.06 | 0.06 | 0.06 | 0.00 | 0.06 | 0.06 | 0.06 | 0.06 | 0.06 | 0.06 | 0.00 |
| | | 20-24 | 0.06 | 0.06 | 0.06 | 0.06 | 0.06 | 0.06 | 0.00 | 0.06 | 0.06 | 0.06 | 0.06 | 0.06 | 0.06 | 0.00 | 0.06 | 0.06 | 0.06 | 0.06 | 0.06 | 0.06 | 0.00 |
| | | 25-34 | 0.06 | 0.06 | 0.06 | 0.06 | 0.06 | 0.06 | 0.00 | 0.06 | 0.06 | 0.06 | 0.06 | 0.06 | 0.06 | 0.00 | 0.06 | 0.06 | 0.06 | 0.06 | 0.06 | 0.06 | 0.00 |
| | | 35-44 | 0.06 | 0.06 | 0.06 | 0.06 | 0.06 | 0.06 | 0.00 | 0.06 | 0.06 | 0.06 | 0.06 | 0.06 | 0.06 | 0.00 | 0.06 | 0.06 | 0.06 | 0.06 | 0.06 | 0.06 | 0.00 |
| | | 45-54 | 0.06 | 0.06 | 0.06 | 0.06 | 0.06 | 0.06 | 0.00 | 0.06 | 0.06 | 0.06 | 0.06 | 0.06 | 0.06 | 0.00 | 0.06 | 0.06 | 0.06 | 0.06 | 0.06 | 0.06 | 0.00 |
| | | 55-74 | 0.06 | 0.06 | 0.06 | 0.06 | 0.06 | 0.06 | 0.00 | 0.06 | 0.06 | 0.06 | 0.06 | 0.06 | 0.06 | 0.00 | 0.06 | 0.06 | 0.06 | 0.06 | 0.06 | 0.06 | 0.00 |
| | | 75-79 | 0.00 | 0.00 | 0.00 | 0.00 | 0.00 | 0.00 | 0.00 | 0.00 | 0.00 | 0.00 | 0.00 | 0.00 | 0.00 | 0.00 | 0.00 | 0.00 | 0.00 | 0.00 | 0.00 | 0.00 | 0.00 |
| | Non-Hispanic White | 15-19 | 0.06 | 0.06 | 0.06 | 0.06 | 0.06 | 0.06 | 0.00 | 0.06 | 0.06 | 0.06 | 0.06 | 0.06 | 0.06 | 0.00 | 0.06 | 0.06 | 0.06 | 0.06 | 0.06 | 0.06 | 0.00 |
| | | 20-24 | 0.06 | 0.06 | 0.06 | 0.06 | 0.06 | 0.06 | 0.00 | 0.06 | 0.06 | 0.06 | 0.06 | 0.06 | 0.06 | 0.00 | 0.06 | 0.06 | 0.06 | 0.06 | 0.06 | 0.06 | 0.00 |
| | | 25-34 | 0.06 | 0.06 | 0.06 | 0.06 | 0.06 | 0.06 | 0.00 | 0.06 | 0.06 | 0.06 | 0.06 | 0.06 | 0.06 | 0.00 | 0.06 | 0.06 | 0.06 | 0.06 | 0.06 | 0.06 | 0.00 |
| | | 35-44 | 0.06 | 0.06 | 0.06 | 0.06 | 0.06 | 0.06 | 0.00 | 0.06 | 0.06 | 0.06 | 0.06 | 0.06 | 0.06 | 0.00 | 0.06 | 0.06 | 0.06 | 0.06 | 0.06 | 0.06 | 0.00 |
| | | 45-54 | 0.06 | 0.06 | 0.06 | 0.06 | 0.06 | 0.06 | 0.00 | 0.06 | 0.06 | 0.06 | 0.06 | 0.06 | 0.06 | 0.00 | 0.06 | 0.06 | 0.06 | 0.06 | 0.06 | 0.06 | 0.00 |
| | | 55-74 | 0.06 | 0.06 | 0.06 | 0.06 | 0.06 | 0.06 | 0.00 | 0.06 | 0.06 | 0.06 | 0.06 | 0.06 | 0.06 | 0.00 | 0.06 | 0.06 | 0.06 | 0.06 | 0.06 | 0.06 | 0.00 |
| | | 75-99 | 0.00 | 0.00 | 0.00 | 0.00 | 0.00 | 0.00 | 0.00 | 0.00 | 0.00 | 0.00 | 0.00 | 0.00 | 0.00 | 0.00 | 0.00 | 0.00 | 0.00 | 0.00 | 0.00 | 0.00 | 0.00 |

*Figure 2: Assortative partnership matrix (left) and uniform partnership matrix (right)*

Under assortative mixing, all calibration targets were satisfied as well as in the empirical mixing, with exception to new diagnoses by race/ethnicity. This is expected as this mixing pattern assumes racial/ethnic groups only mix internally, resulting in increased incidence rates for already burdened groups (such as non-Hispanic Black MSM). For the uniform mixing calibration, all calibration targets are satisfied as well as under the empirical mixing case except we see more new diagnosis among Hispanic MSM when compared to the empirical mixing. Presented below are the calibration plots for new diagnosis by race under assortative and uniform mixing. These are the only plots that show substantial differences from the empirical partnership matrix case shown in section 3.

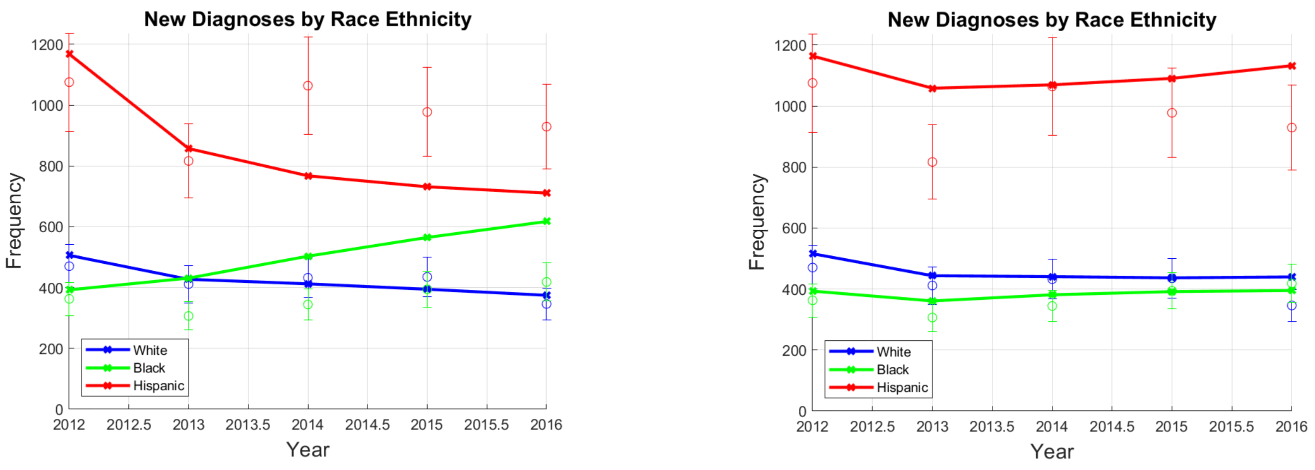

*Figure 3: New Diagnoses calibration under different partnership mixing. Assortative partnership matrix (left) and uniform partnership matrix (right)*

As described in the main manuscript, differences in the magnitude of cumulative infections were observed under different mixing matrices. We attributed a large portion of this to the differences found incidence rate over time in the base case. Presented are the incidence rates, by race, for each mixing pattern over time when no policy is implemented.

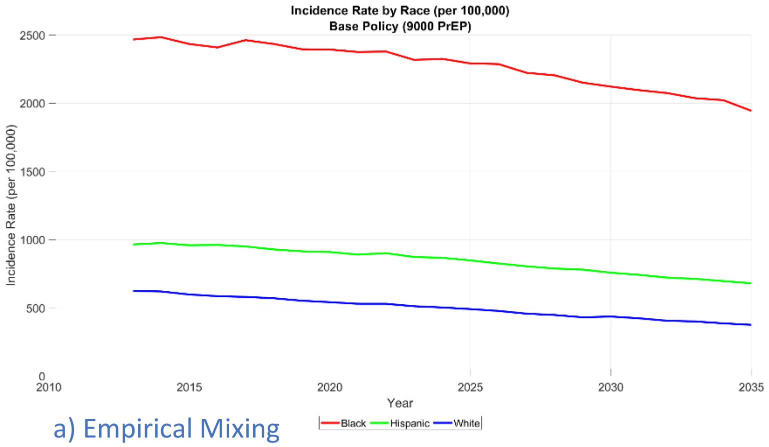

a) Empirical Mixing

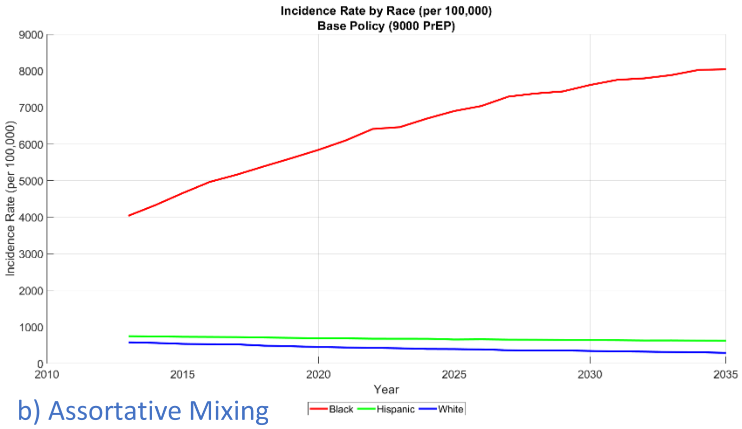

b) Assortative Mixing

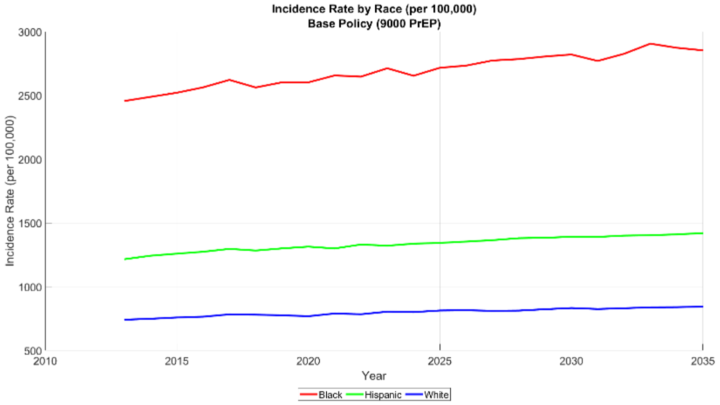

c) Uniform Mixing

*Figure 4: Incidence rate by race, over time, in base case scenario (no allocation policy)*

We also compare the Gini index in 2035 across the three mixing scenarios. We find that the same general trends in Gini index exist regardless of the mixing matrix selected. Individual Gini index values should only be compared to those under the same mixing matrix.

*Table 17: Gini index for three mixing scenarios (1) Empirical Data, (2) Assortative, and (3) Uniform*

| **Policy** | **Empirical Matrix** | **Assortative Matrix** | **Uniform Mixing** |
|---|---|---|---|
| No Intervention | 0.24 | 0.45 | 0.18 |
| Black 3000 | 0.20 | 0.38 | 0.16 |
| Black 6000 | 0.16 | 0.30 | 0.12 |
| Black 9000 | 0.13 | 0.21 | 0.11 |
| Hispanic 3000 | 0.24 | 0.45 | 0.18 |
| Hispanic 6000 | 0.23 | 0.45 | 0.17 |
| Hispanic 9000 | 0.24 | 0.46 | 0.17 |
| White 3000 | 0.25 | 0.46 | 0.19 |
| White 6000 | 0.26 | 0.47 | 0.19 |
| White 9000 | 0.27 | 0.48 | 0.21 |
| Equal 3000 | 0.23 | 0.43 | 0.17 |

| | | | |
|---|---|---|---|
| Equal 6000 | 0.22 | 0.40 | 0.17 |
| Equal 9000 | 0.21 | 0.39 | 0.16 |
| Count 3000 | 0.23 | 0.44 | 0.18 |
| Count 6000 | 0.23 | 0.43 | 0.17 |
| Count 9000 | 0.22 | 0.41 | 0.17 |
| Rate 3000 | 0.22 | 0.41 | 0.16 |
| Rate 6000 | 0.20 | 0.35 | 0.15 |
| Rate 9000 | 0.17 | 0.30 | 0.13 |

#### 9. Incidence Rate and Diagnosis Rate over Time

To reflect how quickly disparities are reduced, we show the incidence rate over time and the new diagnosis rate over time under empirical mixing for Black and Rate policies. These policies at the 9000 PrEP level showcase the largest reduction in disparities. Under the black policy, incidence rates between Black MSM and Hispanic MSM become similar by 2024. Under the rate policy, the Black incidence rate never drops to that of the Hispanic and the benefit drastically diminishes over time. In terms of diagnosis rates, the diagnosis rates drop at a consistent rate until 2029 under the black policy. The decline is relatively constant under the rate policy. Note that a lower diagnosis rate is still a good thing because it reflects fewer HIV positive individuals.

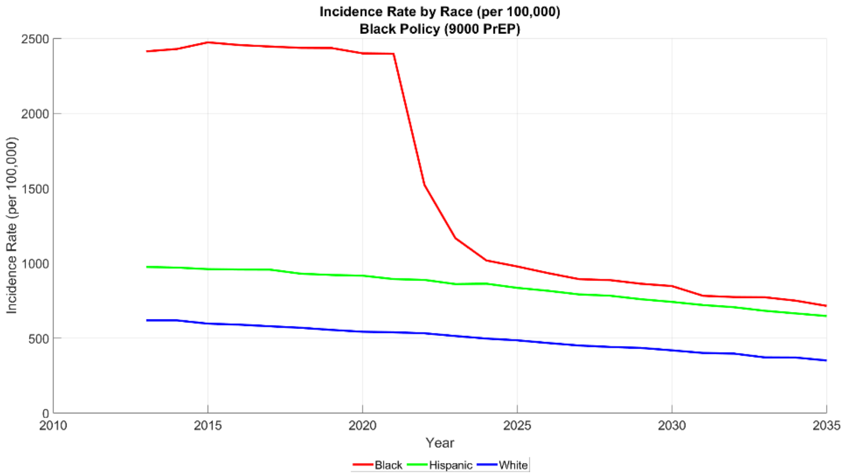

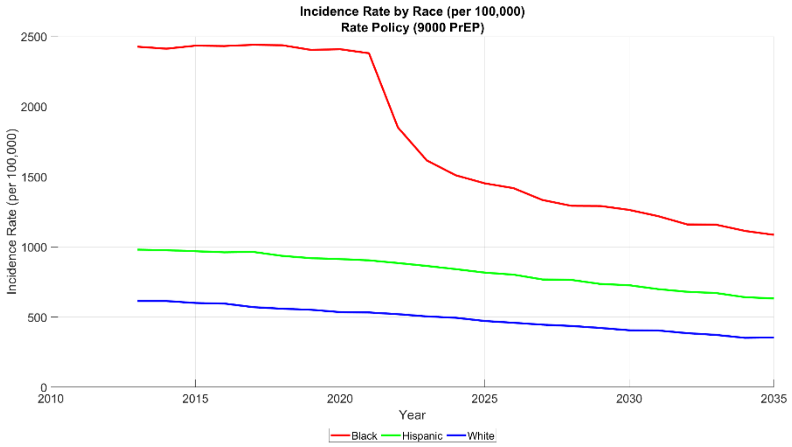

*Figure 5: Incidence rate by race, over time, for the Black and Rate allocation strategies at the 9000 PrEP level*

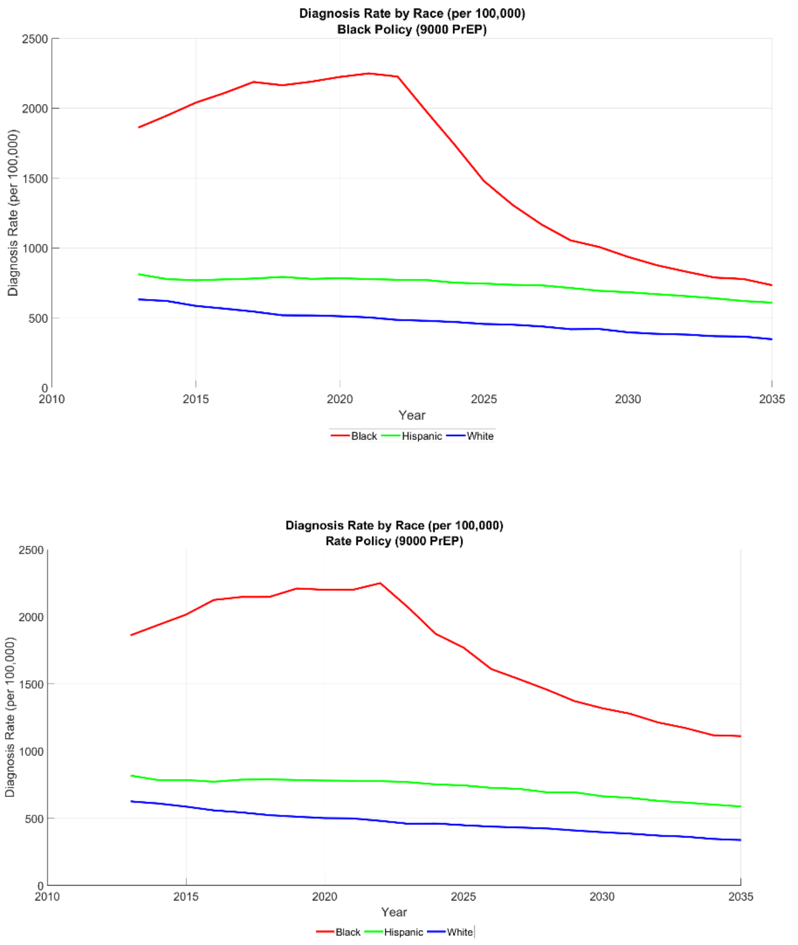

*Figure 6: Diagnosis rate by race, over time, for the Black and Rate allocation strategies at the 9000 PrEP level*

4. United States Census Bureau. QuickFacts: Los Angeles County, https://www.census.gov/quickfacts/fact/table/losangelescountycalifornia/PST045219.

14. Shover C, Javanbakht M, Shoptaw S, et al. High Discontinuation of Pre-Exposure Prophylaxis Within Six Months of Initiation- Los Angeles. In: *CROI*.

15. Shover CL, Shoptaw S, Javanbakht M, et al. Mind the gaps: prescription coverage and HIV incidence among patients receiving pre-exposure prophylaxis from a large federally qualified health center in Los Angeles, California : Mind the Gaps: Cobertura de recetas e incidencia de VIH entre pacientes re. *AIDS Behav* 2019; 23: 2730–2740.

16. Hughes AJ, Mattson CL, Scheer S, et al. Discontinuation of Antiretroviral Therapy Among Adults Receiving HIV Care in the United States. *JAIDS J Acquir Immune Defic Syndr* 2014; 66: 80–89.

17. Centers for Disease Control and Prevention (CDC). Vital Statistics Online, https://www.cdc.gov/nchs/data_access/vitalstatsonline.htm.

18. Kasaie P, Pennington J, Shah MS, et al. The Impact of Preexposure Prophylaxis Among Men Who Have Sex With Men: An Individual-Based Model. *J Acquir Immune Defic Syndr* 2017; 75: 175–183.

19. Koenig LJ, Lyles C, Smith DK. Adherence to antiretroviral medications for HIV pre-exposure prophylaxis: lessons learned from trials and treatment studies. *Am J Prev Med* 2013; 44: S91-8.

20. CDC. *PrEP*, https://www.cdc.gov/hiv/basics/prep.html.

21. Supervie V, Barrett M, Kahn JS, et al. Modeling dynamic interactions between pre-exposure prophylaxis interventions & treatment programs: predicting HIV transmission & resistance. *Sci Rep* 2011; 1: 185.

22. Marshall BDL, Goedel WC, King MRF, et al. Potential effectiveness of long-acting injectable pre-exposure prophylaxis for HIV prevention in men who have sex with men: a modelling study. *Lancet HIV* 2018; 5: e498–e505.

23. Jenness SM, Goodreau SM, Rosenberg E, et al. Impact of the Centers for Disease Control's HIV Preexposure Prophylaxis Guidelines for Men Who Have Sex With Men in the United States. *J Infect Dis* 2016; 214: 1800–1807.

24. Stephens W. PARTNER2 Study: Viral Suppression of HIV Prevents Sexual Transmission Between Gay Men. *AJMC*, https://www.ajmc.com/view/partner2-study-viral-suppression-of-hiv-prevents-sexual-transmission-between-gay-men (2019).

25. LA LGBT Center. Los Angeles LGBT HIV Screening Center Dataset.

32. Centers for Disease Control and Prevention (CDC). *CDC Fact Sheet: HIV Among Gay and Bisexual Men*, https://www.cdc.gov/nchhstp/newsroom/docs/factsheets/cdc-msm-508.pdf.

38. Stats Direct. Gini Coefficeint of Inequality, https://www.statsdirect.com/help/nonparametric_methods/gini_coefficient.htm.

39. Kolm S-C. Unequal Inequalities.I. *J Econ Theory* 1976; 12: 416–422.
